# Müller cell changes and subretinal membrane formation in an eye with multifocal geographic atrophy

**DOI:** 10.64898/2026.01.27.26344802

**Authors:** Malia M. Edwards, D. Scott McLeod, Imran A. Bhutto, Rhonda Grebe, Jeffrey D. Messinger, Andreas Berlin, Shreya Jolly, Autumn M. Knight, Jacques Bijon, K. Bailey Freund, Christine A. Curcio

**Affiliations:** Wilmer Eye Institute, Johns Hopkins University, Baltimore MD, USA; Department of Ophthalmology and Visual Sciences, Heersink School of Medicine, University of Alabama at Birmingham, Birmingham AL, USA; University Hospital Würzburg, Würzburg, Germany; Vitreous Retina Macula Consultants of New York NY, USA; Department of Ophthalmology, New York University Grossman School of Medicine, New York NY, USA

**Keywords:** Müller cells, Age-related macular degeneration, glia, retina, AMD, geographic atrophy, remodelling

## Abstract

**Purpose:** Müller cell (MC) morphology and markers were investigated using histology and immunohistochemistry in an eye with clinically documented multifocal geographic atrophy (GA) and correlated with clinical images.

**Methods:** The donor was followed clinically for five years and last examined six years before death. The superior posterior pole retina was dissected and immunolabeled with antibodies against glial fibrillary acidic protein (GFAP; activated MCs and astrocytes) and glutamine synthetase (GS, MC) and Ulex Europaeus Agglutinin-1 lectin (blood vessels) before embedding for JB-4 cross section analysis. The inferior macula was cryopreserved. Cryosections were immunolabeled with MC homeostatic and activation markers. Transmission electron microscopy (TEM) of the fellow eye was used to study ultrastructure changes.

**Results:** Gross examination demonstrated mottled retinal pigment epithelium (RPE) over presumably calcified drusen. In the submacular retina, MC processes surrounding both drusen and outer retinal pigmented lesions created a large subretinal membrane. Cryosection analysis demonstrated persistence of aquaporin 4 and GS in MCs with both proteins prominently expressed in the subretinal membrane. Increased MC S100B and GFAP expression were also observed in the atrophic area as well as the OJZ. Cryosection labeling and TEM confirmed the MC encasing calcified drusen and RPE debris as well as invading basal laminar deposits.

**Conclusions:** This multifocal GA case demonstrates how MC activation and structural changes surrounding individual drusen could coalesce, contributing to photoreceptor loss. MCs penetrating basal laminar deposits and encasing calcified drusen suggests that they are attempting to clear these and/or protect the retina from harmful contents.

## Introduction

Age-related macular degeneration (AMD) causes irreversible central vision loss among aged adults worldwide.^1^ Dry AMD, the more prevalent form, results in focal loss of photoreceptors, retinal pigment epithelial cells (RPE), and choriocapillaris (CC) in the setting of characteristic extracellular deposits. Advanced dry AMD, or geographic atrophy (GA) can present as either unifocal (one large area of atrophy), or multifocal (several areas of atrophy that coalesce).

Beginning early in AMD progression, Müller cells, the primary glial cell in the retina, become activated overlying drusen, as demonstrated by glial fibrillary acidic protein (GFAP) expression.^2–6^ As the disease progresses, these glial cells become activated and remodel, changing shape and location within the retina. As part of this remodeling process, Müller cells create subretinal glial membranes in eyes with unifocal GA.^2,4,6–8^ However, the Müller cell remodeling, and the mechanisms regulating this, in multifocal GA remain poorly defined.

Here, we consider two non–mutually exclusive mechanistic models to explain Müller glial remodeling in multifocal GA. First, we hypothesize that individual drusen act as localized triggers for Müller cell nuclear repositioning and descent of the external limiting membrane (ELM), generating multiple discrete Müller cell–defined borders that subsequently coalesce into a continuous subretinal glial membrane as atrophy expands. Second, we propose that altered distribution of key Müller cell functional proteins—most notably aquaporin-4 (AQP4) and glutamine synthetase (GS)—reflects a compensatory response to disrupted RPE-mediated metabolic and osmotic homeostasis, rather than a purely degenerative loss of Müller cell function. Consistent with this framework, we recently reported that Müller cells in GA retain expression of essential functional proteins, but with markedly altered cellular domain localization. Most strikingly, the polarized expression of AQP4, a key osmoregulatory protein, is shifts drastically in the atrophic area in eyes with unifocal GA.^9^ Rather than being prominently expressed within endfeet at the inner limiting membrane ILM, AQP4 is expressed diffusely throughout Müller cell processes with the strongest expression within the subretinal glial membrane. Strong AQP4 expression was also noted within Müller cell processes through all retinal layers at the border of atrophy. Müller cells play a key role in the border of atrophy in the neurosensory retina, the descent of the external limiting membrane (ELM) towards Bruch’s membrane.^7,8,10,11^ This border is visible in high quality optical coherence tomography (OCT) and cleanly separates an area of photoreceptor depletion in the atrophic area (inner junctional zone, IJZ) from the surrounding area of potentially salvageable photoreceptors (outer junctional zone, OJZ).

In the present study, we investigated the Müller glial response in an eye with clinically documented multifocal GA using both flatmount and cryosection immunohistochemistry, JB-4 histology and, in the fellow eye exhibiting similar characteristics, using transmission electron microscopy (TEM). In addition to the expression of signature Müller cell functional proteins, vimentin, cellular retinal binding protein (CRALBP) and GS, we also investigated markers of glial activation, GFAP and S100B. S100B is a chemokine that is expressed by astrocytes both in the brain and retina as well as some activated Müller cells.^12–14^ While less studied in the retina, S100B has been studied extensively in relation to stroke and Alzheimer’s disease where it is believed to contribute to inflammation.^15–18^ To our knowledge, this is the first report to demonstrate immunohistochemically the retinal glial changes associated with multifocal GA. A previous report on the fellow eye of this donor addressed histologic correlates of fundus autofluorescence variation, including atrophy initiation at ELM descents over individual drusen.^10^

## Methods

### Human Donor information

Retrospective review of medical records, imaging data and the histopathologic study were approved by institutional review boards of the Manhattan Eye, Ear, and Throat Hospital/Northwell Health, the University of Alabama at Birmingham, and Johns Hopkins University School of Medicine, respectively. This study was conducted in accordance with the Declaration of Helsinki and the Health Insurance Portability and Accountability Act of 1996. Utilization of this human tissue was in accordance with approval of the Joint Committee on Clinical Investigation at JHU. The donor presented herein was a Caucasian female in her early 90s with clinically diagnosed multifocal GA. She was non-smoking donor who suffered from cardiovascular disease, including mitral valve prolapse, systemic hypertension and hypercholesterolemia. Age-matched controls (n=5) with no ocular abnormalities were used to demonstrate the normal expression patterns of Müller cell proteins. The average age of these control donors was 85 years (+/- 4.3 years). The Müller cell protein expression in three of these control donors was previously reported.^9^ Table 1 summarizes donor information.

**Table 1:** Human Donor Eyes.

| Patients | Age Range [years] | Sex | AMD status | Cause of Death |
| --- | --- | --- | --- | --- |
| GA | 90-95 | F | Multifocal GA | Cardiovascular disease |
| Control 1 | 80-85 | M | Normal | Renal failure |
| Control 2 | 80-85 | F | Normal | Cerebrovascular accident; myocardial infarct |
| Control 3 | 85-90 | F | Normal | Alzheimer's Disease |
| Control 4 | 80-85 | M | Normal | Parkinson's disease |
| Control 5 | 90-95 | M | Normal | Rectal Sheath hematoma |

F, female; M, male.

### Donor eye collection and tissue preparation

The GA donor eye was collected 5 hours post mortem by personnel of the Eye-Bank for Sight Restoration (NY) and opened anteriorly to improve fixative penetration.^19^ The right eye (OD) was fixed in 4% paraformaldehyde (PFA) in phosphate buffer overnight and then placed in 1% PFA at 4°C for six months until shipping and processing. The left eye (OS) was preserved in 1% PFA and 2.5% glutaraldehyde and processed for ultrastructural analysis by light and transmission electron microscopy (TEM). The posterior pole of the OD eye was imaged upon arrival at the Wilmer Eye Institute. The central retina was bisected, with the inferior half cryopreserved as a full-thickness eyecup and the superior half processed as separate retinal and choroidal flatmounts. The choroid is reported in a separate paper.^20^ Control eyes were either fixed exactly as the right eye of the GA donor (N=2), at six hours post-mortem, or fixed in 2% PFA in phosphate buffered saline (PBS) containing 5% sucrose within 24 hrs post-mortem (N=3).

### Flatmount Immunohistochemistry

Retinal tissue containing the fovea and optic nerve head was processed for flatmount immunohistochemistry as described ^2,3^ and compared to control posterior pole retinas. Briefly, the retina was blocked in 5% goat serum in tris-buffered saline containing Triton X-100 (TBS-T) and 1% bovine serum albumin (BSA) overnight at 4°C. After washing, the retina was incubated in primary antibody cocktail containing chicken anti-GFAP (Invitrogen, 1:500) and mouse anti-GS (Invitrogen, 1:500) prepared in TBS-T with BSA for 72 hrs at 4°C. Following three washes, retinas were incubated in secondary antibody cocktail containing TBS-T, goat anti-chicken Alexa Fluor 647 (Invitrogen, 1:500), goat anti-mouse cy3 (Jackson Immunoresearch, 1:500) and Fluorescein isothiocyanate (FITC)-conjugated *Ulex Europaeus Agglutinin* 1 (UEA-1) lectin (vascular endothelium; Genetex Inc, Irvine CA, 1:100) for 48 hrs. After final washes, retinas were imaged on a Zeiss 710 confocal microscope. All imaging settings (laser power, gain, etc) were kept the same for control and GA donor eyes. To visualize overall tissue architecture, differential interference contrast (DIC) images were also collected.

### JB-4 embedding and analysis

After imaging, the GA retina was fixed flat by placing between two layers of a Nitex mesh placed in a modified Swinney filter holder, flat fixed in 25% Karnovsky’s fixative^21^ and embedded in glycol methacrylate (JB-4; Polysciences, Inc. Warrington, PA, USA) as previously described.^4^ Two ½ -µm-thick sections were cut using a dry glass knife on a Sorvall MT2-B Microtome (Norwalk, CT, USA). Sections were transferred to drops of water on glass slides and dried on a hot plate prior to staining with periodic acid/Schiff’s (PAS; Sigma-Aldrich, St. Lous, MO, USA) and hematoxylin (Sigma-Aldrich HHS16). Images were captured on a Zeiss Photomicroscope II (Carl Zeiss, Inc. Oberkochen, Germany) using a Gryphax NAOS 20 Megapixel Full HD USB 3.0 Color Digital Microscope Camera (Jenoptik Jena, Germany).

### Cryosection immunohistochemistry

Sections (8 µm thick) were air dried and treated in −20°C methanol for 5 minutes. After air drying, sections were blocked in 10% goat serum prepared in TBS with 0.1% BSA for 1 hr and then incubated in primary antibody cocktail (Table 2) prepared in TBS with 0.1% BSA for 2 hrs. Sections were washed in TBS and incubated in secondary antibody cocktail prepared in TBS containing DAPI for chromatin (Invitrogen, 1:1000) and UEA lectin (Genetex 1:100) for 30 min. Finally, tissue autofluorescence was quenched by treating sections in Sudan black B (1% freshly prepared in 70% ethanol; Electron Microscopy Sciences; Hatfield, PA USA) for 10 minutes in the dark followed by washing in distilled water. All steps were carried out at room temperature. Images were captured on a Zeiss 710 confocal microscope and processed using Zen software. Images were collected with a 20x objective with or without tiling. All image settings (laser power and gain) were kept the same for control and GA donor eyes for each antibody combination. DIC images were also collected.

**Table 2:**
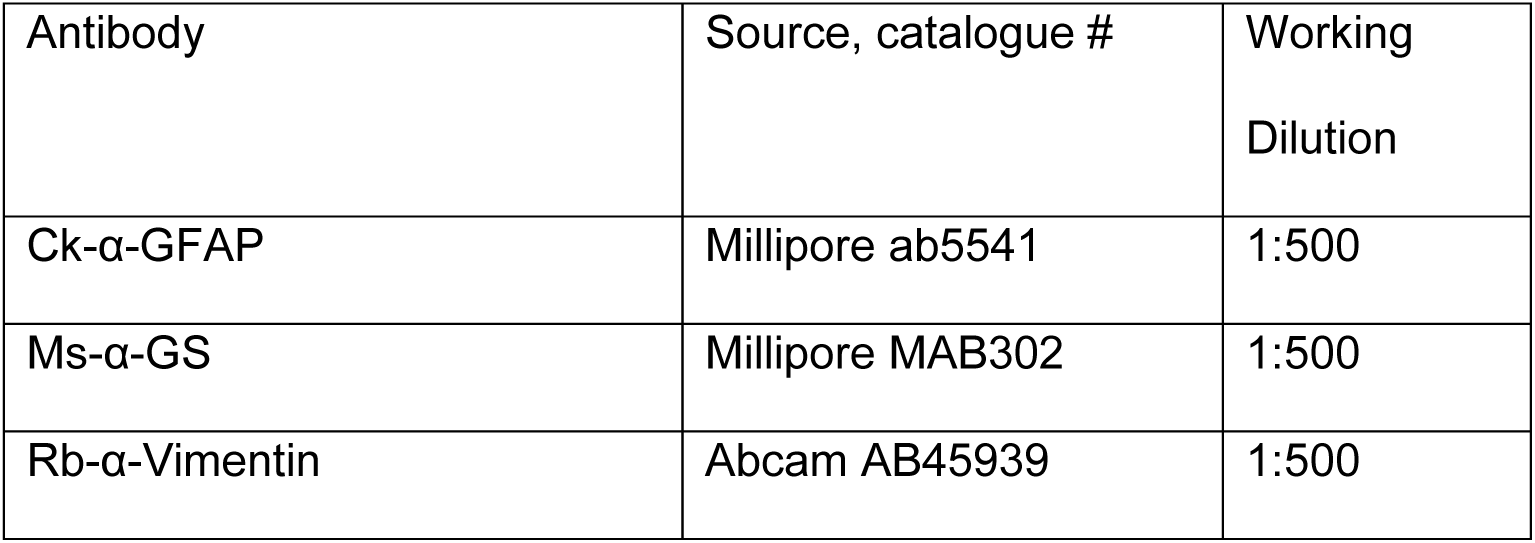
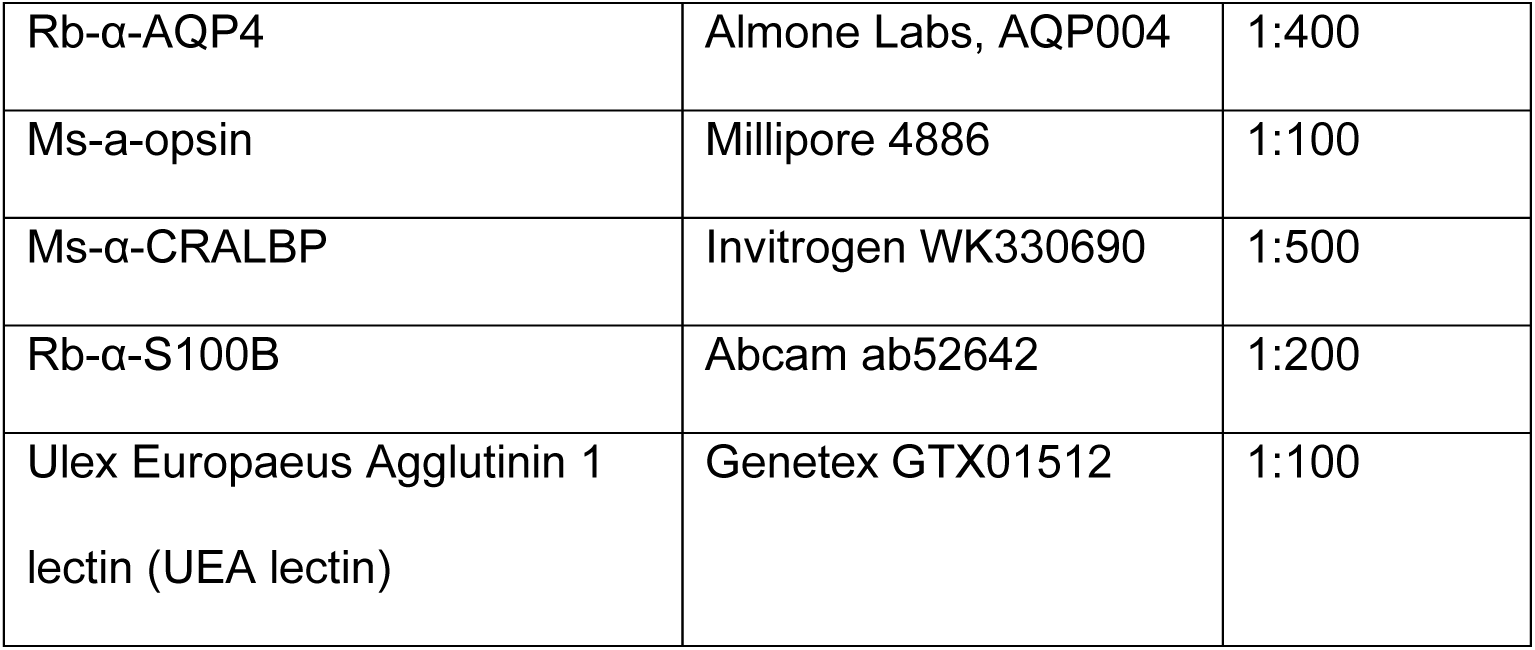
Primary antibodies used in this study. Ms: Mouse, Rb: Rabbit, Ck: Chicken

### Transmission Electron Microscopy (TEM)

The fellow eye was fixed, processed and imaged as described.^10^ Sections from this eye were prepared for TEM. For correlative high-resolution histology and electron microscopy, a rectangular tissue block (8 × 12-mm wide) containing the fovea and optic nerve was postfixed in 1% osmium - tannic acid - paraphenylenediamine and embedded in epoxy resin (Polybed 812 resin kit, Electron Microscopy Services, Hatfield, PA). Submicrometer sections (∼0.8 µm thick) were cut with a histology grade diamond knife (Diatome Histo knife, Electron Microscopy Services, Hatfield, PA) on an ultramicrotome (Ultracut, Leica) and saved at 30- to 60-μm intervals on glass slides. Interleaved slabs approximately 30-µm-thick were saved for TEM. Selected slabs were re-embedded by first flattening them between two microscope slides on a low temperature hot plate. Under stereoscopic magnification, the flattened section was compared to the image of submicrometer section. A 2-mm-wide region of interest was dissected out with a sharp scalpel (#11; Graham Field Sterile Ophthalmology Micro Scalpels 2979#45). This region was placed along the vertical surface of an embedding mold (Pelco Flat embedding mold, Ted Pella Catalog #105, Fisher Scientific) and propped up vertically by a new blank resin blank. New resin was added, and the mold was heated at 75C° for 18 hours. Sections were cut at gold thickness on a Leica UC7 ultramicrotome employing a Diatome “Ultra” series diamond knife and placed onto carbon coated slotted copper grids (2×1 mm) or uncoated 100 mesh hexagonal copper grids (Electron Microscopy Services). Sections were stained with 2% uranyl acetate and post stained with Reynolds lead citrate. Images were taken on a H7600 Hitachi TEM (Hitachi High-Technologies Corp., Tokyo, Japan) equipped with an Advanced Microscopy Techniques model XR80, 2KCCD model (Woburn, MA, USA).

## Results

### Clinical history and fundus presentation

The donor had been followed clinically over the course of 6 years and was last seen 5 years prior to death. The donor had undergone bilateral cataract removal with lens implantation. Fundus photography (Fig. 1A) demonstrated drusen in the macular and mid-peripheral regions. Fundus autofluorescence revealed atrophic areas with RPE loss (Fig. 1B). A small area with exudative choroidal neovascularization (CNV) was observed superior to the optic disc at baseline (Fig. 1B arrows) and followed up. There was no exudation in the last clinical exam. Multiple large, calcified drusen as well as central outer retinal and RPE atrophy were evident on the OCT B-scan at the last visit (Fig. 1C, D). The choroidal vessels also appeared abnormally dilated (Fig. 1D). Gross images taken prior to removing the retina demonstrated the presence of drusen in the affected macula (Fig. 1E, F).

**Figure 1.**
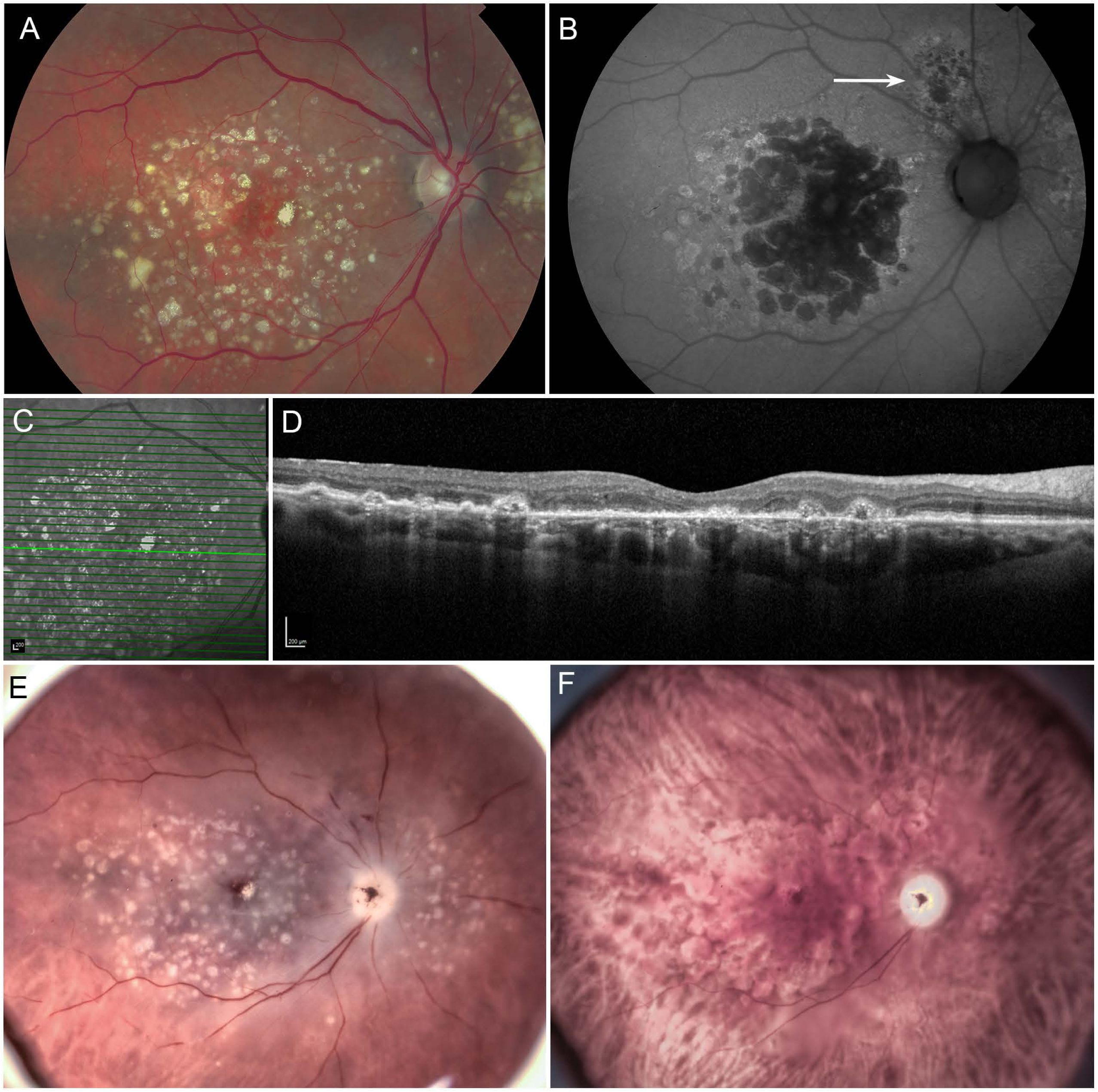
Multimodal clinical and laboratory imaging. The last clinical images of the multifocal GA donor eye were collected 5 yrs prior to her death. (A) Fundus photography shows drusen in the macula and perimacula. (B) Fundus autofluorescence revealed the atrophic area with RPE loss as well as a small area with exudative choroidal neovascularization (CNV) superior to the optic disc (arrow). (C, D) The last OCT-B scan revealed multiple large, calcified drusen as well as central outer retinal and RPE atrophy. The choroidal vessels also appeared abnormally dilated. (E, F) Widefield ex vivo imaging with the retina in place demonstrated drusen in the affected macula. Scale bars indicate 200 µM (C & D).

### Flatmount analysis revealed a large discontinuous subretinal membrane created by Müller cells encasing drusen occupying atrophic areas within the posterior pole

The control retinas, when viewed from the side of the ELM, were unremarkable at low magnification (Supplemental Fig. 1A). The honeycomb-like pattern created by Müller cell processes at the ELM were evident at high magnification (Supplemental Fig. 1B). In images of the GA eye, the atrophic areas were well demarcated. In these regions, numerous long, thin processes, positive for GS and GFAP, extended along the outer retinal surface (Fig. 2A). These processes were interrupted frequently by calcified drusen, many of which remained adherent to the retina during dissection from the choroid (Fig. 2, 3). These drusen were enveloped by Müller cell processes (Fig. 2B-D). The density and complexity of these glial processes was best demonstrated at higher magnification and could be segregated by z slices at different focal planes (Fig. 3A-F). Multiple Müller cell processes, positive for both GS and GFAP, extended through the ELM to completely envelope some drusen (Fig. 3B). Between drusen, processes from Müller cells extended horizontally along the outer retinal surface (Fig. 3). In the focal plane containing the ELM, a disrupted ELM was evident with prominent GS staining in areas lacking drusen (Fig. 3C). Müller cell processes also appeared to both penetrate and encircle other drusen often segregating individual calcified nodules (Fig. 3C-E). Müller cell processes extended along the subretinal surface to fill the gaps created by drusen in the ELM focal plane (Fig. 3F). Therefore, the Müller cells appeared to sequester the retina from drusen components.

**Figure 2.**
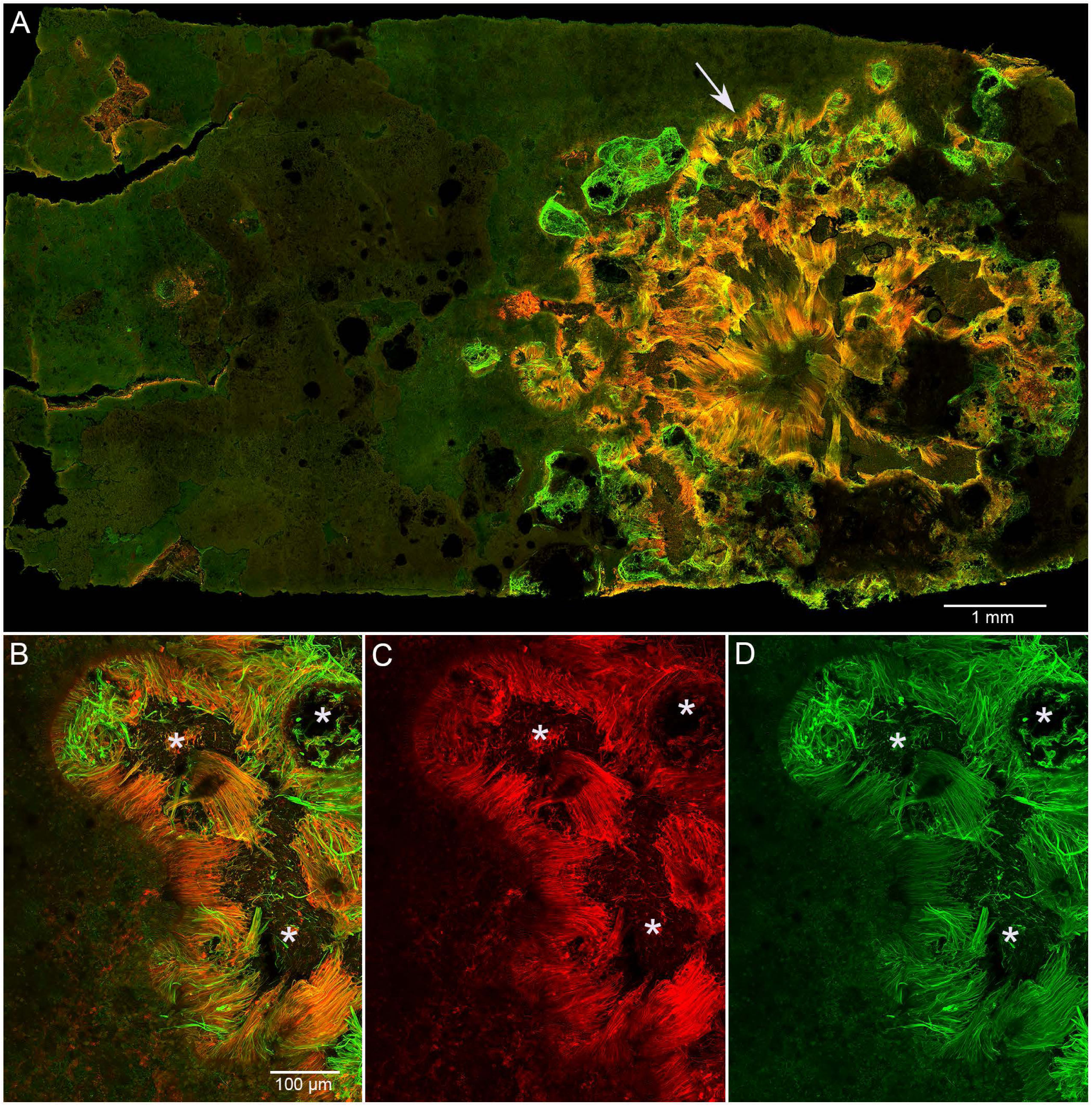
Activated glia viewed from underneath the multifocal GA retina. (A) Half of the macular region stained with GFAP (green) and GS (red). A glial membrane was observed on the outer retinal surface (arrow). The glial processes within the membrane were positive for both GFAP and GS. In some areas, Müller cell processes appeared to be surrounding drusen. Higher magnification more clearly shows Müller cell processes, positive for both GS and GFAP, extending over areas previously occupied by drusen mechanically lost during dissection (asterisks). Scale bars indicate 1mm (A), 100 µm (C-D).

**Figure 3.**
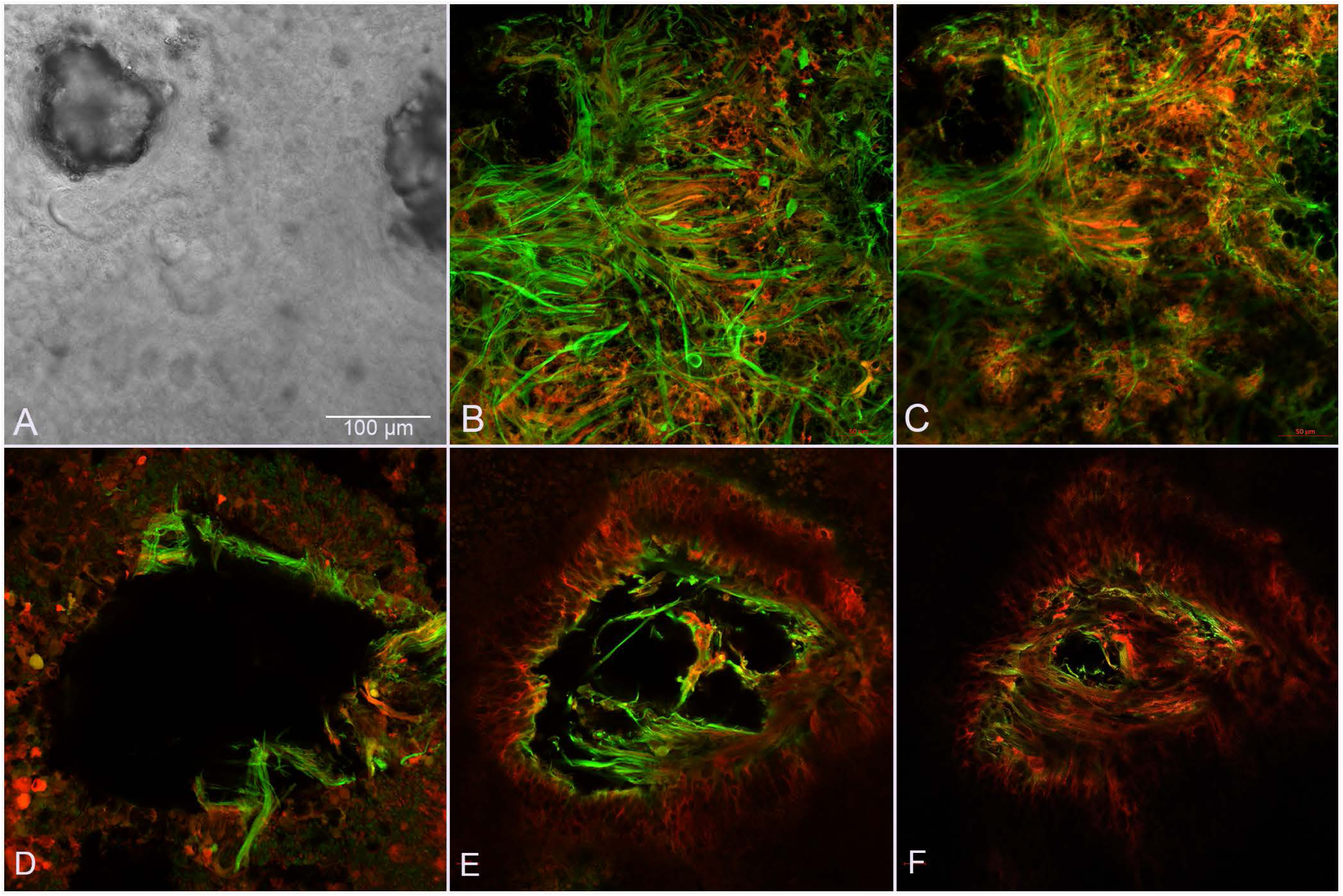
High magnification Z series of multifocal GA retinal flatmount stained with GFAP and GS. (A) DIC imaging of the retinal flatmount shows two drusen in the upper right and left corners. (B) In the focal plane at the level of the outer retina, Müller cell processes positive for GFAP and GS extended horizontally along the outer retinal surface. In the upper left corner, Müller cell processes created a thick band surrounding the drusen. On the upper right, processes were covering the druse. (C) In a focal plane closer to the ELM, the disorganized ELM was obvious with intense GS labeling. Processes were present surrounding the drusen on the upper left with some appearing to extend into individual nodules within the druse. The druse on the upper right contained visible nodules which were each encircled by GFAP and GS-positive processes. (E) In another area, in the focal plane most external to the retina, multiple Müller cell processes, positive for GFAP and GS, surrounded a large druse. On one side, a wall-like structure was created with Müller cell processes extending over the druse. (F) In a focal plane closer to the ELM, Müller cell processes entering the druse were even more evident. (G) Müller cell processes covered almost the entire gap created by the druse at the level of the ELM. The honeycomb-like pattern of the ELM is evident on either side of the druse. Müller cell processes covering the druse extended horizontally across the retinal surface creating swirls. Scale bars indicate 100 µm.

In one area highlighted in Figure 4, a RPE-capped druse remained adherent to retina and was evident in the gross photograph. This same area was visualized in the flatmount showing Müller cell processes extending onto the surface to ensheathe the druse (Fig. 4B). JB-4 sections of the same area, stained with PAS and hematoxylin, revealed a thick basal laminar deposit (BLamD) overlying the druse (Fig. 4 C-E). Glial processes were observed surrounding the BLamD and the druse (Fig. 4D, E). Melanin containing organelles were observed within Müller cell processes adjacent to the drusen and BLamD (Fig. 4E).

**Figure 4.**
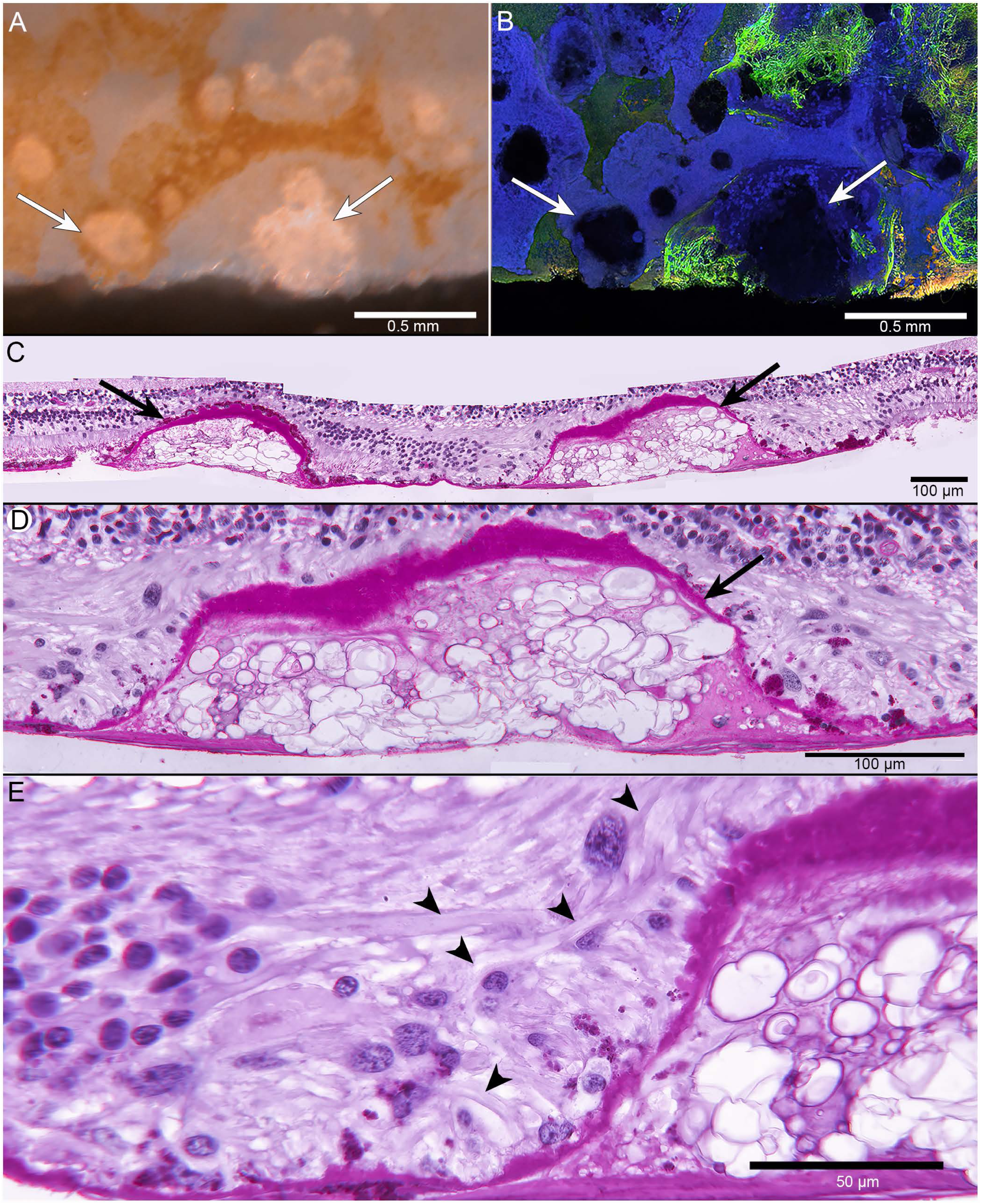
Müller cell association with drusen in en face and cross-sectional view. (A) Gross photographs of the multifocal GA donor eye showed drusen (arrows) surrounded by RPE. (B) The flatmount retina stained with GFAP (green), GS (red) and UEA lectin (blue), and imaged with the ELM en face, shows the same drusen (arrows) which remained attached to retina during dissection. RPE were nonspecifically stained with UEA lectin. Müller cell processes were observed overlying drusen as well as between drusen. (C-E) JB-4 sections of the flatmount retina were stained with PAS/hematoxylin. The drusen with multiple nodules were clearly visible as was the BLamD overlying the drusen and in adjacent areas (arrows). Müller cell processes were observed surrounding the drusen and were disorganized (arrowheads in E). Scale bars indicate: A, B: 0.5 mm; C, D: 100 µm and E 50 µm.

### Analysis of cross-sections further demonstrates the glial membrane and its complexity as well as Müller cells ensheathing drusen

In the control retina, GFAP immunoreactivity was confined to astrocytes in the nerve fiber layer and isolated Müller cell processes (Supplemental fig 2). CRALBP labeled entire cell body and processes of Müller cells as well as RPE cells (Supplemental Fig 2). Within the atrophic region of the GA eye, CRALBP expression appeared slightly reduced compared to that in the control eye in the inner retina but was still present. It was more prominent external to the outer plexiform layer, where photoreceptors were absent (Fig. 5A, B). Müller cell processes, positive for CRALBP and GFAP, were disorganized throughout the retina, having lost their usual linear structure (Fig. 5A-C). As seen in flatmounts, these processes extended through the outer retina, defined herein as external to the outer plexiform layer regardless of whether photoreceptors were present, and directly contacted drusen (Fig. 5A-D). These drusen created mounds, forcing the layers above to adapt to the new topography. Dense bands of GFAP and CRALBP double-positive processes were observed over drusen and on Bruch’s membrane.

**Figure 5.**
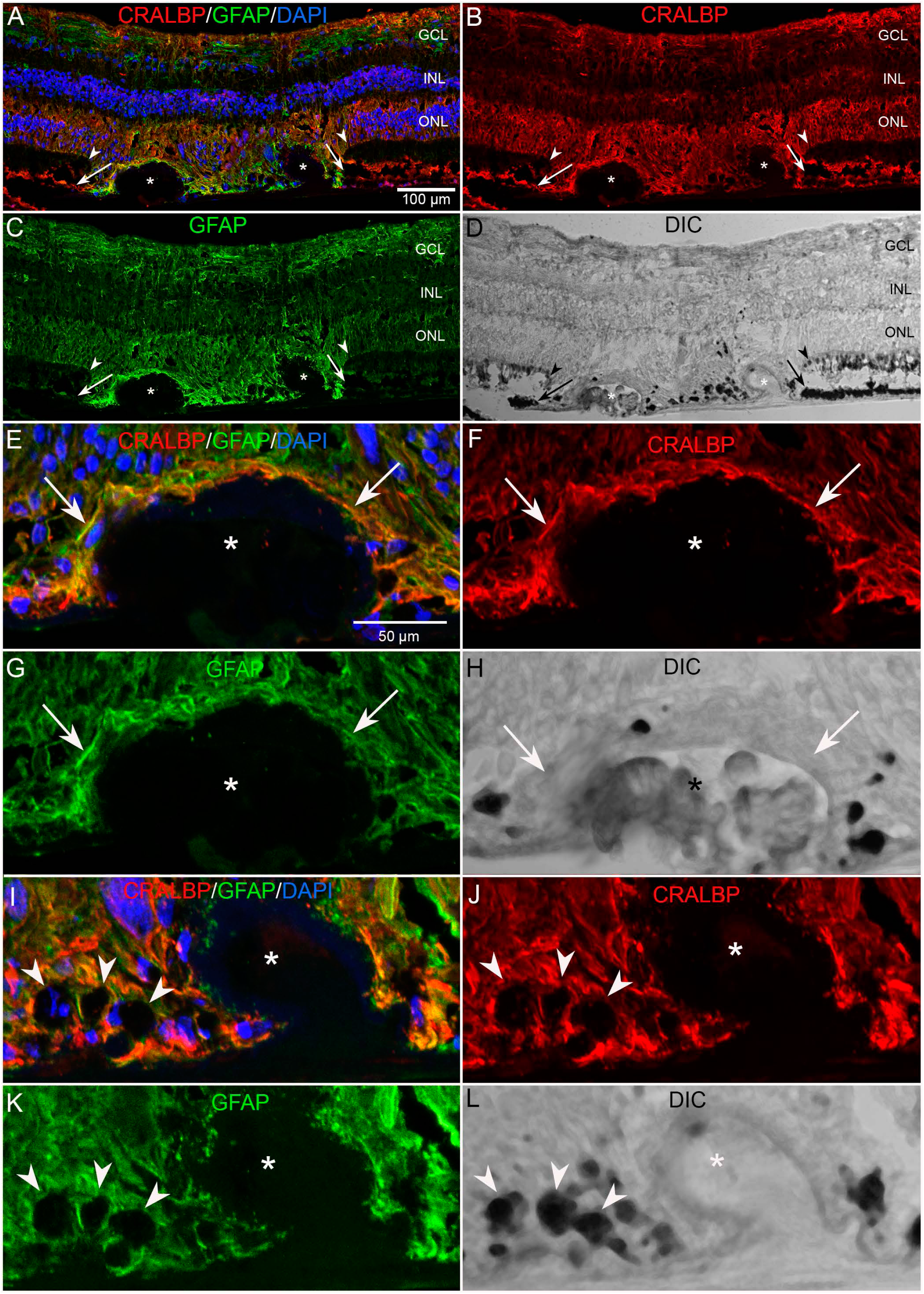
Gliosis and retinoid markers of retinal cross sections from the multifocal GA donor eye. (A) Within the affected, Müller cells were positive for both GFAP (green) and CRALBP (red). DAPI (blue) demonstrates presumed repositioning and dislocation of Müller cell nuclei around drusen and in between two druse (asterisks). Müller cells, positive for both GFAP and CRALBP created the ELM descent (arrowheads) on either side of drusen. (A-C) At the border of atrophy (arrow), RPE cells were also positive for CRALBP. (D) DIC imaging demonstrated pigmented cells, presumably RPE, at the base as well as in between the two calcified drusen. (E-H) A dense band of double-positive processes were observed over the larger drusen (asterisk) with a BLamD (arrows). (I-L) In between drusen, pigmented cells (arrowheads) were surrounded by GFAP/CRALBP-positive Müller cell processes arrowheads. Asterisks indicate calcified drusen in all images. All drusen in this image were devoid of RPE. Scale bars indicate: 100 µm (A-D), 50 µm (E-L). GCL: ganglion cell layer, INL: inner nuclear layer, ONL: outer nuclear layer.

Some drusen were observed in the affected area with photoreceptors and RPE still present (Fig. 5). In these areas, Müller cells created an ELM descent on either side of drusen when RPE were still present between two drusen (Fig. 5A-D, arrowhead). On the non-atrophic aspect of these ELM descents (OJZ), the outer nuclear layer (ONL) had fewer nuclei but similar thickness compared to unaffected areas. Nuclei, which appear to be within GFAP and CRALBP positive Müller cell processes, were observed on the outer retinal surface on both sides of the drusen as well as between two drusen. A dense line of double-positive processes was also observed running perpendicular to Bruch’s membrane (Fig. 5 E-H). Pigmented cells, presumably of RPE origin but lacking CRALBP signal, were noted on the retinal aspect of this glial membrane (Fig. 5). These cells were often observed mounding on top of one another, particularly adjacent to drusen (Fig. 5, G-L). In several areas, nuclei were also observed in the inner and outer plexiform layers, suggesting that cells were migrating throughout the retina as would occur with remodeling.

### Müller cells exhibited increased S100B and GFAP expression in the atrophic retina

In control retinas and in unaffected regions of the posterior pole within the GA eye, S100B and GFAP immunoreactivity were confined to astrocytes and isolated Müller cell processes (Supplemental Fig. 2). In the affected macular area, however, Müller cells strongly expressed both GFAP and S100B (Fig. 6A-C). Their processes occupied locations both internal and external to persistent BLamD overlying calcified drusen. In some cases, Müller cells processes extended into the druse itself. Throughout the atrophic area, S100B expression within Müller cells was most prominent in the outer retina and in processes extending subretinally (Fig. 6A, B). As was observed with other markers, S100B and GFAP-positive cells/processes were located at the base of each druse. While Müller cells normally have a vertical and linear morphology, this usual arrangement was disrupted by the presence of numerous drusen in the affected areas. Some Müller cells and their processes within the subretinal membrane appeared to lie horizontally along a BLamD or directly in contact with Bruch’s membrane. Between drusen, mounds of pigmented cells were often observed in the outer retina, surrounded by GFAP and S100B-positive Müller cell processes (Fig. 6).

**Figure 6.**
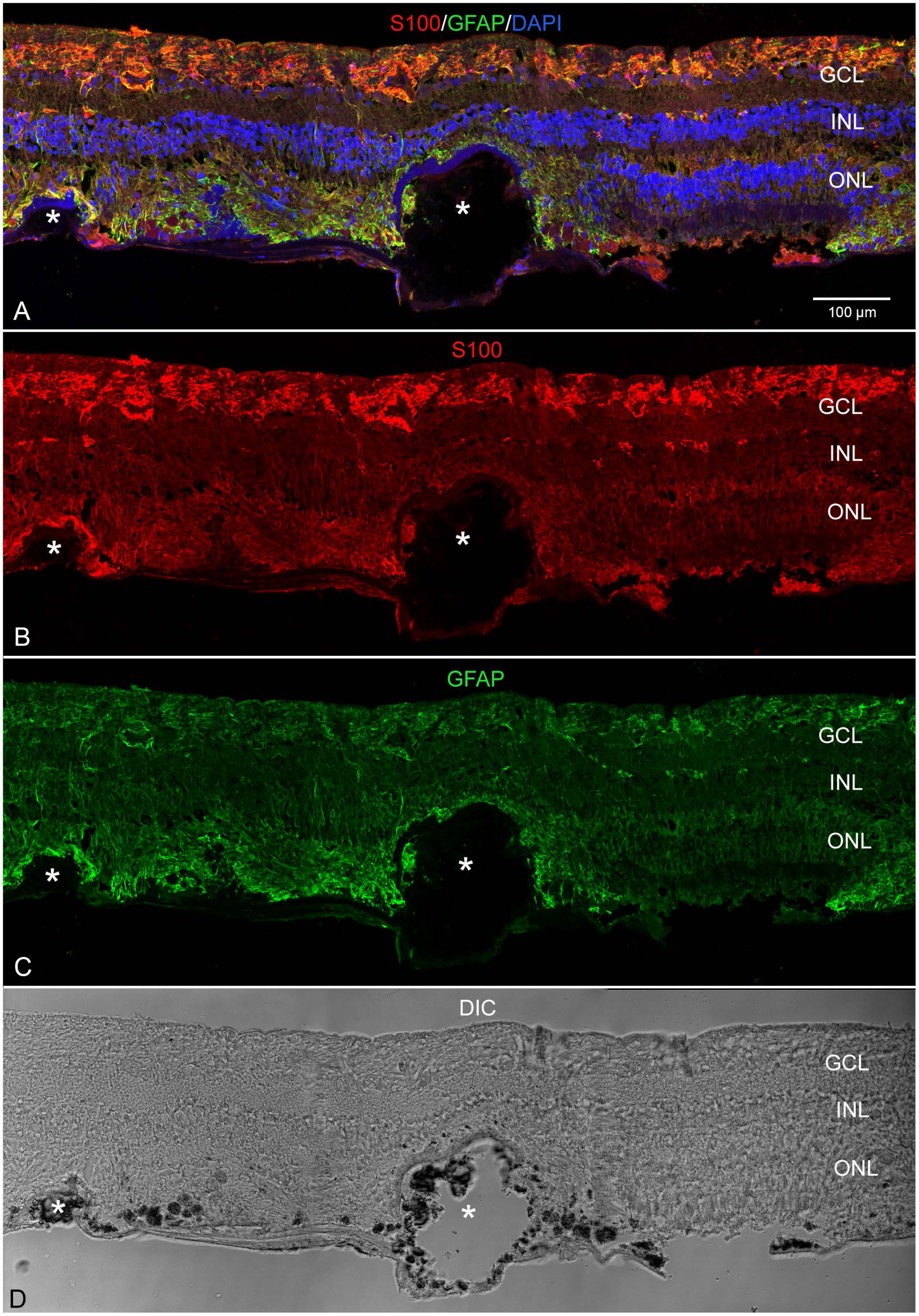
Müller cells were activated and enwrapped calcified drusen in the affected area of the GA eye. (A) S100B (red), GFAP (green) and DAPI (blue) staining through the pathologic area in the case eye. S100B and GFAP-positive processes were also observed above and below a BLamD over two apparently calcified drusen. Müller cell processes overlying drusen had projections into the drusen. A very thin layer of GFAP and S100B-positive glial cells was also observed on the Bruch’s membrane side of the BLamD. (B) As in the control areas, S100B was prominently expressed by astrocytes, however, strong expression was also observed in Müller cell processes. (C) GFAP was expressed by astrocytes and activated Müller cells. (D) DIC microscopy demonstrated pigmented cells, presumably RPE aggregates, mounding on one another within the retina. Asterisks indicate drusen. Scale bar indicates 100 µm. GCL: ganglion cell layer, INL: inner nuclear layer, ONL: outer nuclear layer.

### Müller cell expression of AQP4 shifted in the atrophic area while GS expression persisted

In age-matched control retinas, AQP4 was confined to astrocytes and Müller cell endfeet at the inner limiting membrane as well as perivascular Müller cell processes in the deep capillary plexus (Supplemental Fig. 2). Little, if any, AQP4 is observed at the ELM in control retinas. In the unaffected areas of the GA posterior pole, GS and AQP4 expression patterns were similar to that observed in controls (data not shown). In the non-atrophic aspect of the ELM descent (OJZ), GS immunostaining was more intense in the outer retina, including Henle’s fiber layer, compared to the inner retina (Fig. 7A, C). Aquaporin 4 was prominently expressed in the inner retina and perivascular Müller cells but also diffusely expressed in their radial processes throughout the retina (Fig. 7A, B). At the ELM descent, AQP4 immunostaining was increased within Müller cell processes throughout all retinal layers (Fig. 7A, B; arrow). Within the IJZ, the atrophic aspect of the ELM descent, Müller cells showed intense staining for AQP4 but very low, if any, GS immunoreactivity (Fig. 7A-C arrow). Directly adjacent to this, within the more atrophic region, GS expression was prominent in the outer retina. Müller cells, positive for AQP4 and GS, were overlying and encapsulating calcified drusen, similar to observations made with other markers. The expression of these proteins, however, was distinct around drusen, GS being more pronounced on the Bruch’s membrane aspect of drusen while AQP4 was more prominent on the inner aspect (Fig. 7 A-C). Particularly strong expression was noted in the perivascular processes (paired arrows in Fig. 7A-D) as well as in the subretinal glial membrane (Fig. 7A, B). In the atrophic area, GS was slightly reduced in the inner retina compared to controls but increased in the outer retina and subretinal membrane (Fig. 7E-G). Consistent with other Müller cell markers, a continuous thin layer of GS and AQP4-positive cell processes with a few scattered, nuclei were present underlying the BLamD. A thick banded structure was observed with DIC imaging between the retina and thin glial membrane (Fig. 7H). Pigmented cells were again noted on the inner aspect of the glial membrane. Here, they were apparently encased by AQP4 and GS-positive Müller cell processes.

**Figure 7.**
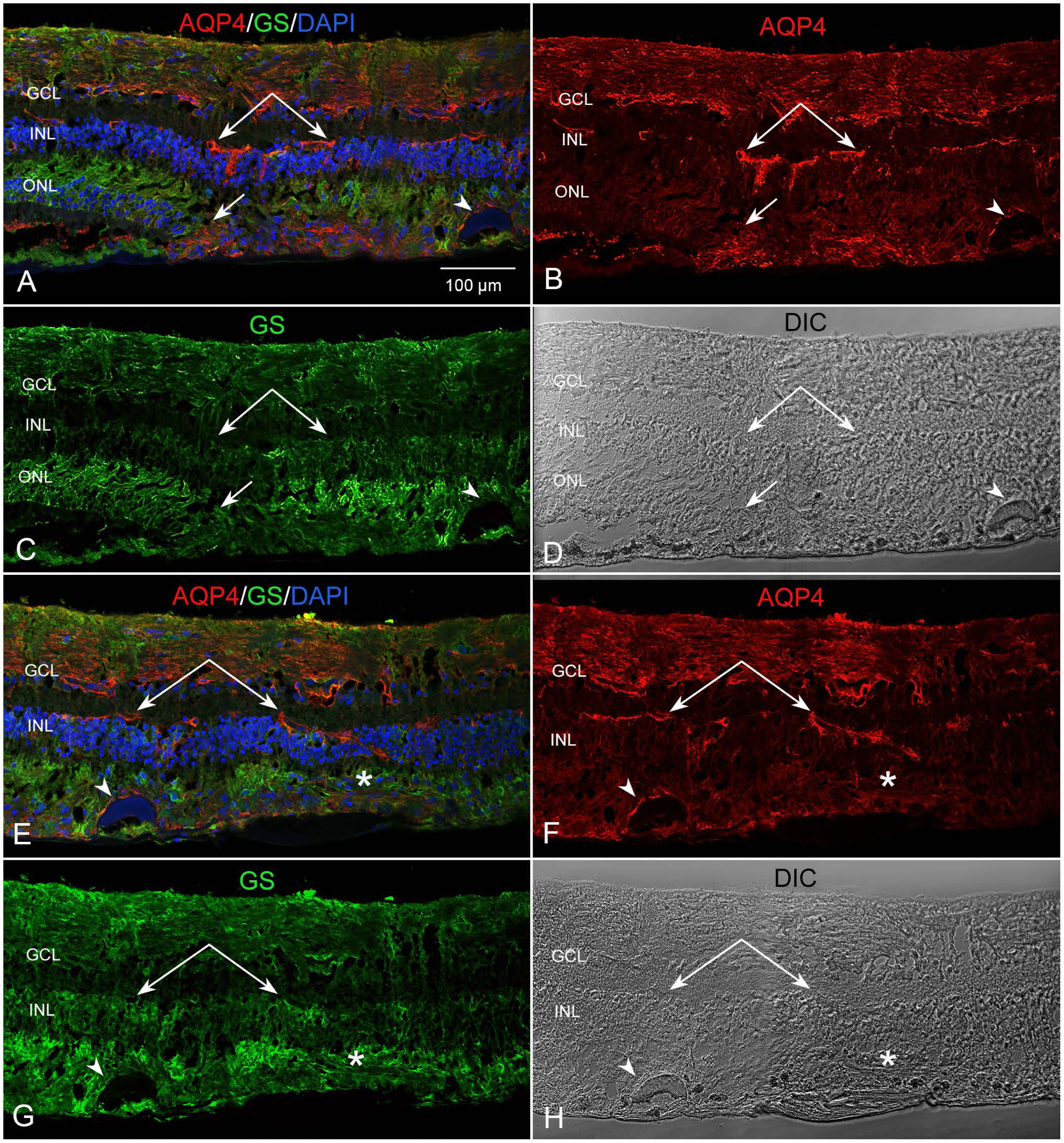
Müller cell expression of GS and AQP4 was altered in the affected area, viewed in cross section. (A) GS (green), AQP4 (red) and DAPI (blue) staining at the atrophic border. (B) In the OJZ, AQP4 was localized to astrocytes and Müller cell endfeet in the nerve fiber as well as perivascular processes (paired arrows) with some diffuse staining in the radial processes. In the atrophic area, AQP4 was also observed in the subretinal glial membrane and in the IJZ. (C) GS was observed in Müller cell radial processes on both the inner and outer junctional zones adjacent to the ELM descent (arrow). Reduced GS expression was observed in the outer retina within the OJZ. Some background autofluorescence of the RPE was observed in the 568 wavelength (green channel). (D) DIC imaging showed the drusen which was surrounded by Müller cell processes. (E) In the adjacent atrophic area, the ONL was replaced by a glial membrane. (F) AQP4 was expressed diffusely in Müller cell processes. Particularly intense staining was noted over drusen (arrowhead), in perivascular processes and in the subretinal membrane. (G) GS staining remained in Müller cell processes even in the atrophic area and in the glial membrane. (H) DIC imaging shows drusen and BLamD. Scale bars indicate 100 µm. Paired arrows indicate perivascular processes positive for AQP4. Arrowheads indicate small drusen with thick BLamD. All drusen in the imaged areas were calcified and lacked RPE. Asterisks indicate the glial membrane in E-H. GCL: ganglion cell layer, INL: inner nuclear layer, ONL: outer nuclear layer.

### Vimentin and opsin staining demonstrated the Müller cell structural changes

Vimentin labeled the entire length of Müller cells in controls and non-pathologic regions of the GA retina (Supplemental Fig. 3). Opsin was confined to the cone outer segments. At the atrophic border in the GA retina, vimentin-positive Müller cell processes formed the ELM descent (Fig. 8A, C). In the OJZ, Müller cell processes within the Henle fiber layer were slightly thicker than normal, indicative of gliosis. In the IJZ and atrophic area, vimentin-positive Müller cell processes were no longer linear but rather extended through the retina at different angles and into the subretinal space. As noted with other markers, a very thin layer of vimentin-positive processes with a few nuclei was also observed under the BLamD (Fig. 8A, C, G). Opsin was observed in the outer segments. Inner segments were shortened or absent. Mis-localized somatic opsin staining was also observed in cells within Henle fiber layer (Fig. 8). Similar to observations with other antibodies, pigmented cells, presumably RPE or debris from fragmented RPE cells, was embedded within the vimentin-positive membrane. These were separated from the BLamD by Müller cell processes. Importantly, these pigmented cells did not express vimentin, which is a marker of epithelial mesenchymal transition (EMT).

**Figure 8.**
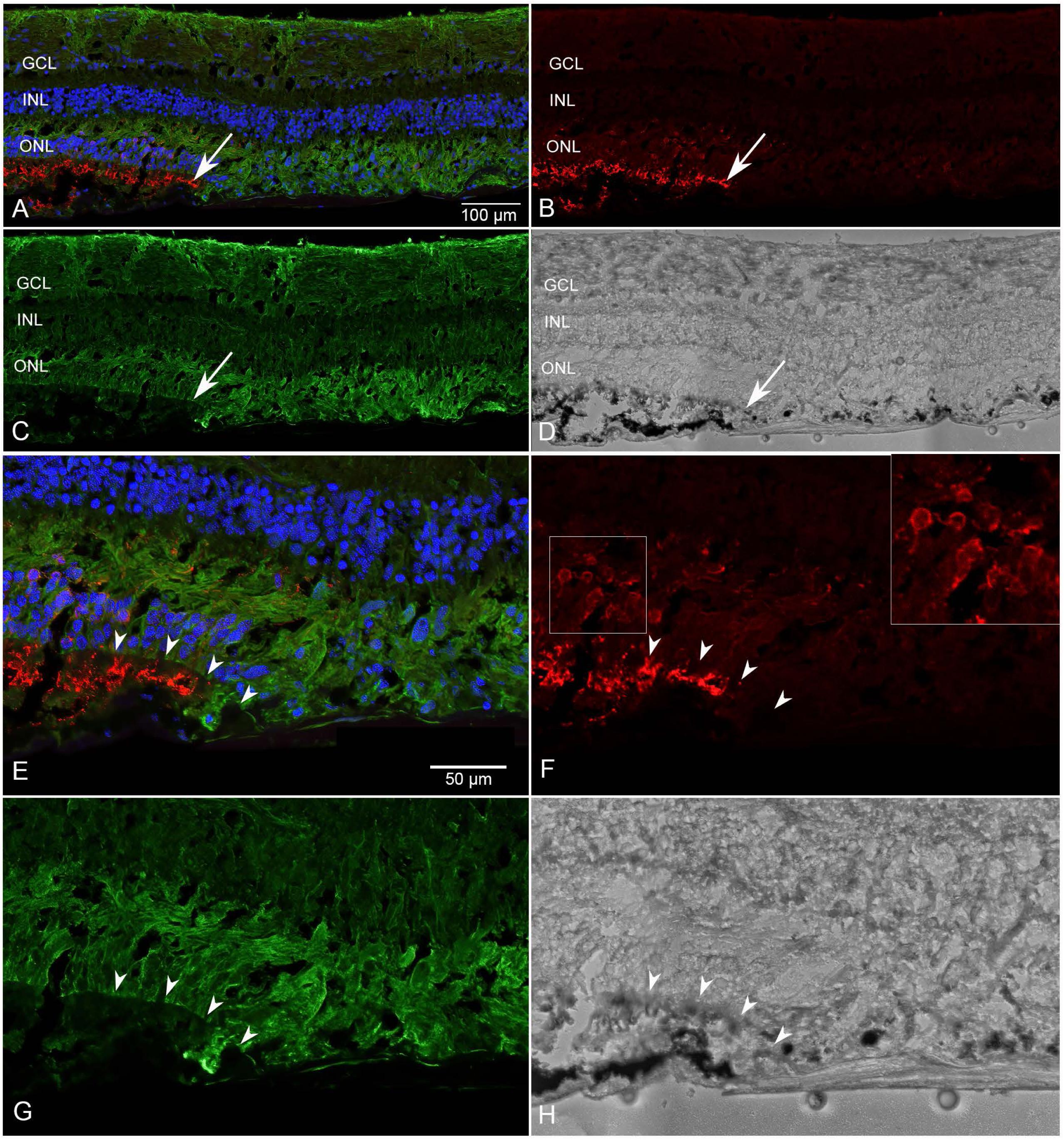
Müller cell structural changes and redistribution of opsin in the outer junctional zone. (A-C) At the border in the GA donor eye, vimentin-positive Müller cells (green) created the ELM descent (arrow). LM opsin-positive inner segments (red) were observed only in the OJZ. Mislocalized somatic opsin was observed within the outer nuclear layer and Henle’s Fiber Layer adjacent to the ELM descent. (D) DIC imaging revealed BLamD in the atrophic retina as well as pigmented cells within the vimentin-positive glial membrane. (E-H) High magnification images more clearly demonstrated the vimentin-positive ELM (arrowheads) and Müller cells extending into the subretinal space creating the ELM descent. (F) Somatic opsin mislocalization was also better demonstrated at higher magnification. (H) DIC imaging showed RPE intact on the nonatrophic aspect of the ELM descent (arrowheads). A BLamD was also observed in the atrophic area below the glial cells. Pigmented cells were embedded in the vimentin-positive glial membrane. Importantly, these pigmented cells did not express vimentin, which is an EMT marker. GCL: ganglion cell layer, INL: inner nuclear layer, ONL: outer nuclear layer.

### Glial cell processes invade BLamD and drusen

As mentioned above, in atrophic areas where RPE were lost or only isolated RPE remained, Müller cell processes ensheathed individual drusen. This was better observed at higher magnification (Fig. 9A-D). Here, BLamD was also evident over calcified drusen. In these cases, Müller cells created two distinct lamellas, one overlying the BLamD and another beneath the drusen, separating these structures from Bruch’s membrane. Müller cell processes, positive for GFAP, were observed penetrating the BLamD as well as enveloping individual hydroxyapatite nodules^22^ within the drusen (Fig. 9A-D, E). Toluidine blue staining and TEM of the fellow eye confirmed Müller cell processes surrounding drusen (Fig. 9F-H). In addition, finger-like processes were noted extending from horizontally oriented Müller cells in the subretinal space. These penetrated into BLamD (Fig. 9G, H).

**Figure 9.**
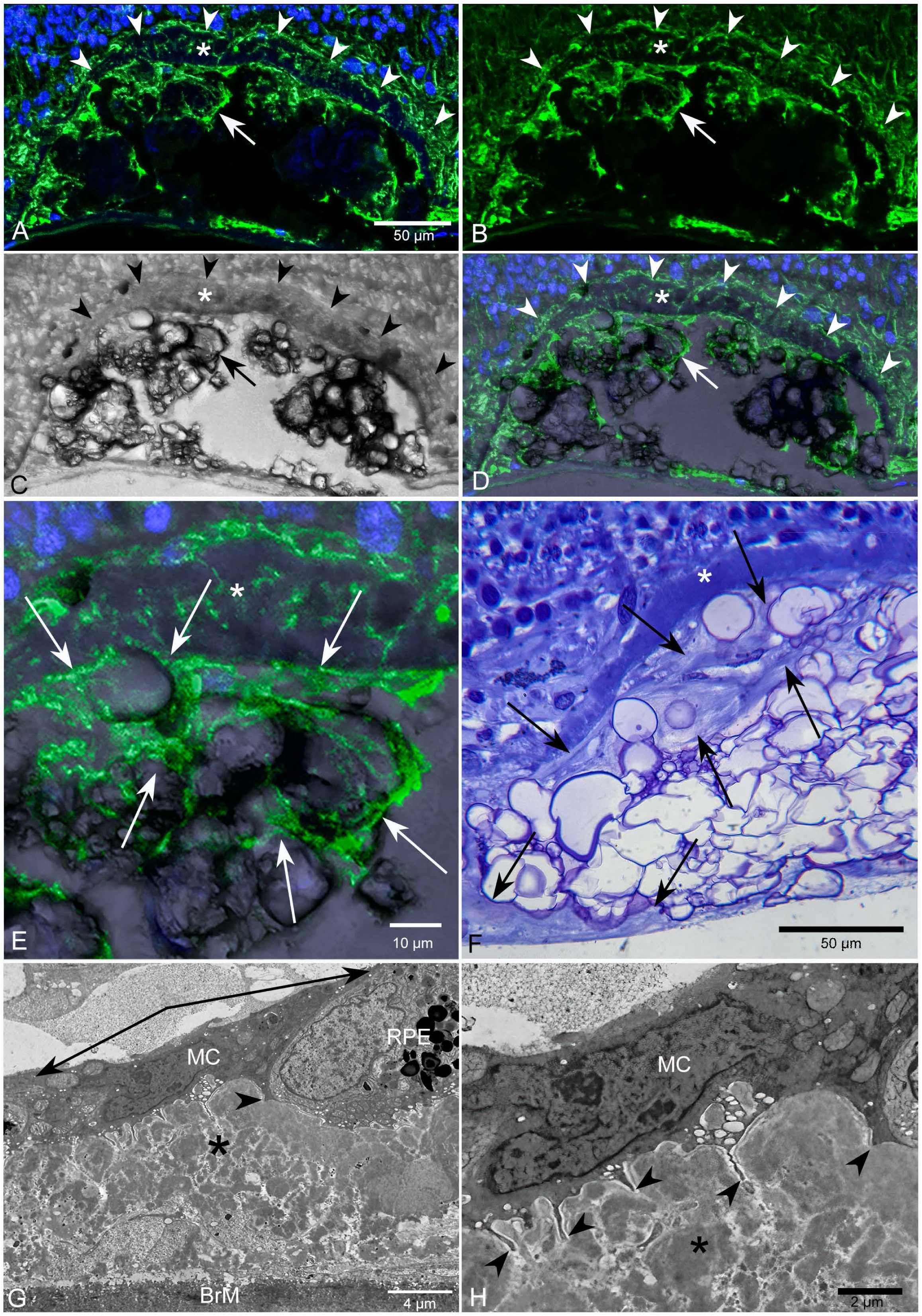
GFAP staining shows Müller cell enveloping drusen. (A-E) A cross section from the GA donor eye stained with GFAP (green) and DAPI (blue) focusing on a large druse with persistent BLamD and lacking RPE. (B) Two thick lines of GFAP stained processes were observed overlying the drusen. GFAP processes were also observed within the druse while small, finger-like Müller cell processes penetrated the BLamD covering the druse. (C) DIC imaging revealed the nodules within the druse as well as the BLamD. (D) Imaging of the staining along with DIC. (E) Higher magnification more clearly showed the GFAP-positive processes encasing hypoxyapatite nodules of the druse (arrows). (F) Semi-thin sections of the partner eye stained with toluidine blue revealed a BLamD as well as calcified drusen with apparent Müller cell processes. (G) Transmission electron microscopy also demonstrated Müller cell (MC) processes overlying BLamD (asterisk) and RPE (arrows). (H) Finger-like processes, resembling those observed with GFAP staining, were noted extending from horizontally oriented Müller cells in the subretinal space with TEM. Scale bars indicate: A-D, F: 50 µm, E: 10 µm, G: 4 µm, and H: 2 µm. MC: Müller cells; BrM: Bruch’s membrane;

### Müller cells over drusen demonstrate features of ELM descent formation

In the unaffected macular retina, as in the control, S100B and GFAP were primarily confined to astrocytes. When drusen was present, however, both proteins were also expressed in overlying and neighboring Müller cell radial processes (Fig. 10A-D). While Müller cell processes did not extend beyond the ELM in these areas, isolated nuclei within GFAP and S100B positive cell processes were observed just below the ELM. Numerous nuclei in the INL and ONL were observed between layers, suggesting their repositioning within the retina. This reduced the outer plexiform layer thickness and created a concave appearance. RPE, while dysmorphic, were still present except at the innermost aspect of the druse, where photoreceptor segments were missing (Fig. 10D). Müller cell remodeling over individual drusen with RPE was also evident with vimentin staining (Fig. 10E, G). Vimentin-positive cells also appeared to extend beyond the ELM along the outermost aspect of drusen. Opsin-positive outer segments were also lost at the apex of drusen and reduced along the edges (Fig. 10E, F).

**Figure 10.**
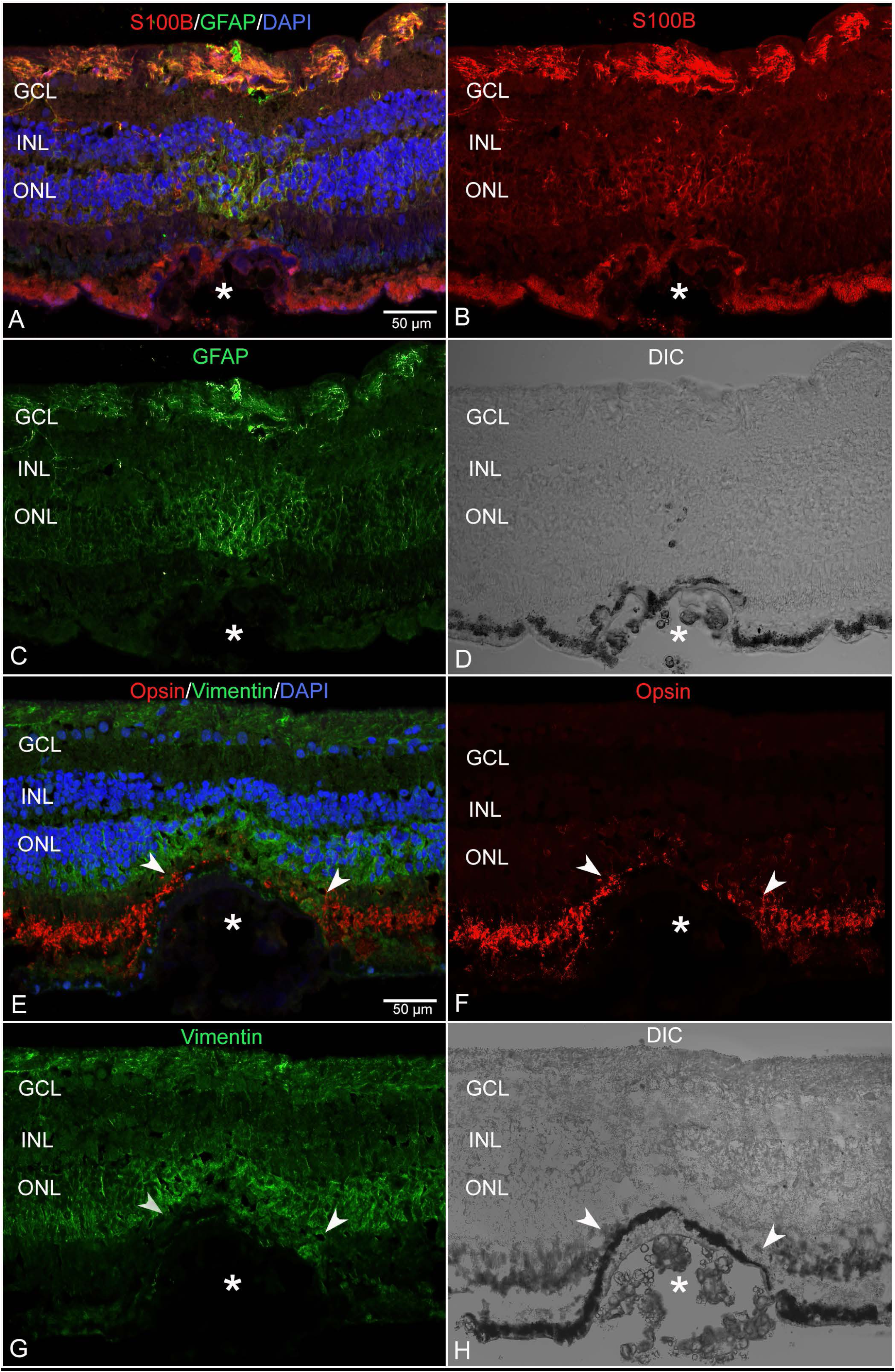
Glial markers in nonatrophic retina with RPE-capped partially calcified drusen. (A-C) Within the macula, but outside the atrophic area, S100B (red) and GFAP (green) were primarily confined to astrocytes. Some background autofluorescence of the RPE was observed in the 568 wavelength (red channel) due to insufficient Sudan black quenching. Both proteins were also expressed in Müller cell processes directly above drusen (asterisk). The Müller cell processes here lost their linear morphology and were instead oriented towards the top of the druse. (D) DIC imaging demonstrated intact RPE which became thin at the inner aspect of the druse while photoreceptor inner and outer segments were absent. (E-G) Vimentin (green) and opsin (red) staining of cryosections from the multifocal GA donor eye demonstrated Müller cell remodeling surrounding drusen. On either side of the druse, vimentin-positive cells were observed extending beyond the ELM (arrowheads). Opsin-positive inner segments were shortened or missing overlying the apex of the druse (asterisk) but some mislocalized opsin was observed within the retina. (H) DIC imaging revealed RPE intact except at the top of the druse. Scale bars indicate 50 µm. GCL: ganglion cell layer, INL: inner nuclear layer, ONL: outer nuclear layer.

## Discussion

Although previous reports have included donor eyes with multifocal GA,^23,24^ these studies did not separate unifocal and multifocal GA in their analysis. Therefore, this is the first comprehensive histological report of Müller cell changes in a retina with multifocal GA. As in unifocal GA, we observed an extensive Müller cell subretinal membrane occupying the atrophic areas in this donor eye. Overall, the changes in Müller cell protein expression were similar to those we recently reported in eyes with focal GA.^9^ As will be discussed below, the multi-lobular nature of the atrophy, however, also created some unique features compared to more focal GA. These could represent differences between these two forms of GA or could be due to different states of severity as there were more surviving RPE present in the multifocal donor eye. The multifocal atrophic lesions in this eye created many distinct individual boundaries between atrophic and non-atrophic areas, including islands and peninsulas trapped between atrophic spots that initiated atop drusen.^10^ These presented numerous areas to observe ELM descents and associated changes in Müller cell activity and protein expression.

### The Müller cells ensheathing drusen create a subretinal membrane

With regards to the glial membrane, the multifocal nature of this atrophy created a more disrupted or disjointed membrane than those observed in unifocal GA..^2,4,7,8^ Rather than one large confluent membrane, GFAP and GS-positive processes completely ensheathed calcified drusen which separated from Bruch’s membrane and adhered to the retina in the flatmounts. This occurred both in areas with RPE and photoreceptor degeneration and in adjacent areas where these cells were present. Cross-sectional analysis confirmed that some Müller cell processes penetrated the calcified drusen and/or were overlying BLamD, while others extended along the outer retinal surface, separating these deposits from Bruch’s membrane.

The complexity of the Müller cells with their extensive intertwined processes made distinguishing individual cells nearly impossible yet confocal microscopy led to insights of cell behavior. Based on the nuclei present and observations of Sox9-positive nuclei in unifocal GA within the subretinal space (unpublished data), we speculate that at least some Müller cells reposition subretinally and lie horizontally along the outer retinal surface, between drusen and the inner collagenous layer of Bruch’s membrane. This was observed in both cryosection immunohistochemistry with multiple Müller cell markers (GS, vimentin, GFAP, S100B) as well as with TEM analysis. Frequent interruptions created by drusen gave the membrane a less intricate appearance than those in unifocal GA which are more dense and multi-layered.^2,4,7,8^ In the most severely affected areas, however, the subretinal glial membrane in this donor was also multi-layered. As these areas also had nearly total RPE and photoreceptor loss,^10^ the differences may represent stages of glial membrane formation corresponding to increased disease severity. Overall, more surviving RPE were present in this multifocal GA eye within the affected macular area compared to unifocal GA eyes we have investigated where RPE were completely atrophic in a large continuous area.^2,4^ While we suspect the RPE in the multifocal GA eye were unhealthy, as signified by the presence of drusen and the discontinuity of the monolayer, they were still present. Similar observations were made in the fellow eye.^10^ Interestingly, photoreceptors were missing or distorted over drusen even when RPE were present, as also seen in the fellow eye.

One other notable difference between this eye and eyes with unifocal GA that we have studied,^2,4^ was reduced adhesion between the retina and choroid during dissection for flatmount preparation. Although the retina was more adherent to the choroid than control eyes, it was not as strongly adherent as in eyes with unifocal GA. In addition, the retina in cryosections detached from the choroid even in atrophic areas in this multifocal GA eye. By contrast, in eyes with unifocal GA, we have found the retina to be strongly adherent to the choroid in atrophic areas of cryosections.^2,4^ We believe this adhesion is due to the Müller cell processes penetrating Bruch’s membrane into the inner choroid.^2,4,7,8,10,25^ In this donor, however, Müller cells appeared to extend horizontally along the BLamD or Bruch’s membrane rather than penetrating through Bruch’s membrane. It is possible that the composition of BLamD and Bruch’s membrane create good substrates for Müller extension so they do not penetrate the choroid. It is also possible that the choroidal structure is different in this donor, preventing Müller cell invasion. While a separate study has been devoted to choroidal pathologic changes,^20^ it is worth nothing that the inner choroidal stroma (intercapillary pillars) exhibited severe hyalinization in this eye. This could have impeded Müller cell processes from invading the choroidal stroma, contributing to reduced adhesion.

The Müller cell membrane’s complexity may create a stronger barrier than is created by tight junctions in the normal ELM. Further research on the permeability of the glial membrane is required to verify this. The glial membrane may be protecting the retina from harmful material in the subretinal space. At the same time, however, it could hinder treatment efficacy. This membrane could also impact the flow of nutrients and waste between the retina and choroid.

The discontinuous membrane in this donor with multifocal GA may create less of a barrier to small molecules or drug therapies compared to membranes in eyes with unifocal GA.

### Müller cell remodeling and retinal homeostasis: important considerations for disease progression and treatment potential

Müller cell remodeling in response to photoreceptor loss has been studied in both animal models and human donor eyes with retinitis pigmentosa as well as AMD.^2,4,26–30^ The Müller cell response to photoreceptor and RPE loss, although secondary to the initial insult, likely impacts retinal homeostasis and treatment efficacy. Given the crucial roles Müller cells play in maintaining retinal homeostasis (ion and pH regulation and osmolarity, etc), we must understand how Müller cell functions are impacted by remodeling and membrane formation as occurs in GA. While Müller cells in this multifocal GA eye retained expression of key Müller cell proteins, AQP4, GS and CRALBP, the localization of these was altered in the atrophic area.

Most strikingly, as we recently reported in unifocal GA eyes and a rat model using subretinal sodium iodate to achieve atrophy bounded by ELM descents^9^, AQP4 polarity was disrupted. While perivascular AQP4 expression persisted, staining increased throughout the Müller cell radial processes and was even stronger in the subretinal membrane. This spatial shift in expression likely compensates for reduced debris clearance and osmoregulation by unhealthy or lost RPE. While this shift in AQP4 is likely beneficial for surviving photoreceptors and other neurons, the increased expression throughout Müller cell processes could negatively impact Müller cell osmoregulation within the retina. This could lead to edema within the retina and/or impaired ability of Müller cells to respond to osmotic stress.

We also observed a slight shift in Müller cell GS expression, resulting in increased GS expression within subretinal Müller cells/processes and reduced expression in the nerve fiber layer. The persistence of GS is consistent with our recent report in eyes with unifocal GA and the rat subretinal sodium iodate model. A greater reduction in GS expression within processes was observed, however, in this multifocal GA eye.^9^ Our data on GS differs from a previous report which observed GS loss in eyes with various stages of AMD.^27^ The previous report, however, used immuno-electron microscopy with a different antibody than reported herein.

Therefore, the discrepancy between our studies is likely due to these technical differences. Additionally, the previous report included both neovascular and atrophic AMD at different stages so the differences could also be due to different disease states.

### Müller cell activation and increased S100B expression could contribute to inflammation in AMD

The expression of both GFAP and S100B within Müller cells in the atrophic retina in the OJZ is not surprising as several reports have demonstrated their activation in AMD.^2,4,6,11,31^ Of particular interest in this study was the focally increased expression over solitary drusen in the non-atrophic retina. S100B is a chemokine which exerts both intracellular and extracellular functions. Müller cells and astrocytes can secrete S100B, contributing to increased extracellular concentration. While nanomolar concentrations are neuroprotective, in micromolar concentrations S100B exerts pro-inflammatory and neurotoxic effects.^13,14^ Müller cells may also secrete S100B into the vitreous and aqueous humor, creating a marker of activation. S100B’s role as a chemokine has led to extensive studies regarding its role in Alzheimer’s disease and other neurodegenerative disorders.^17,32,33^ Given the increase noted with isolated drusen outside the atrophic area, Müller cell S100B expression could contribute to the inflammatory response in AMD. Further research is required to determine when in AMD this increase occurs as well as how this contributes to AMD pathology.

### Individual drusen provide clues regarding ELM descent and membrane formation

While it seems reasonable to assume that the glial membrane forms by Müller cells extending processes into the subretinal space, individual drusen suggest an additional scenario. Over each druse, presumed Müller cell nuclei were observed repositioning into the ONL and beyond the ELM even in areas where photoreceptors and RPE were present. This suggests that Müller cell bodies actually migrate to create the ELM descent, and possibly the subretinal membrane, rather than just extending processes. We previously reported ELM descents over individual drusen in non-atrophic areas and have postulated that atrophy begins when Müller glia permanently fill a gap in the RPE monolayer.^10,34^ The retinal remodeling over drusen create an unusual environment where Müller cells and RPE can directly interact, which is not normally the case. As drusen increase in number and size, Müller cells and processes surrounding these extend horizontally along the subretinal surface towards one another. These individual areas eventually coalesce to create an adhesive membrane. This can be seen in our flatmount preparation (Fig. 3). Importantly, the glial membrane extending between drusen may lie beneath photoreceptors and even RPE that are intact, separating them from Bruch’s membrane and the choroid.

### Müller cells and migrating RPE

Another interesting observation in this case was pigmented cells and debris, likely of RPE origin, that had migrated into the retina seemingly along or within Müller cell processes. These pigmented cells, presumably RPE, were present as far inwardly as the INL. The idea that Müller cells are facilitating RPE migration through the retina toward retinal vessels has been suggested previously.^35–37^ Interestingly, these pigmented cells lacked CRALBP expression, supporting the idea that RPE undergo molecular transdifferentiation, possibly EMT, in AMD.^38,39^ It is important to note, however, that these cells did not express vimentin, as one would expect for cells undergoing EMT. One other possibility is that Müller cells, microglia or macrophages have digested dead RPE and are channeling them through the retina. Unfortunately, our antibodies for microglia (Iba1) or macrophages (CD68) did not work well on these sections, perhaps due to the length of fixation.

### Study limitations

One limitation of the present study is that it is based on a single set of donor eyes. Therefore, one must consider the influence that other health issues, including vascular disease, could have exerted on the retinal health. We have reduced the influence of these factors by looking at sections outside the atrophic area, often within the same section in the macula as well as outside the macula, for comparison. Moreover, our results regarding Müller cell remodeling are consistent with those obtained from eyes with unifocal GA that we have investigated. Our data are significant for providing molecular information about the status of cells that are increasingly visible through multimodal OCT-anchored clinical imaging. Thus, a natural history of what we describe in histology may be determined with precision, in vivo.

## Data Availability

All data produced in the present study are available upon reasonable request to the authors

## Acknowledgements

The authors are grateful to the eye donors and their families without whom this work would not be possible. The authors also thank Yonejung Yoon, MSc, PhD, of the Eye-Bank for Sight Restoration (NYC) for timely retrieval of donor eyes as well as Dylan Shin, Olivia Wanex and Maryangeles Vasquez for assistance with immunohistochemistry.

## Abbreviations

GA: geographic atrophy
GFAP: glial fibrillary acidic protein;
GS: glutamine synthetase;
UEA-1: Ulex Europeaus Agglutinin 1
TEM: Transmission electron microscopy
RPE: retinal pigment epithelium
AMD: Age-related macular degeneration
CC: choriocapillaris;
ELM: external limiting membrane;
AQP4: aquaporin-4
OCT: optical coherence tomography
OJZ: outer junctional zone
IJZ: inner junctional zone
CRALBP: cellular retinal binding protein
PFA: paraformaldehyde
PBS: phosphate buffered saline
TBS-T: Triton X-100;
BSA: bovine serum albumin;
FITC: Fluorescein isothiocyanate;
DIC: differential interference contrast;
PAS: periodic acid/Schiff’s;
CNV: choroidal neovascularization;
BLamD: basal laminar deposit;
ONL: outer nuclear layer;
INL: inner nuclear layer;
EMT: epithelial mesenchymal transition;
GCL: ganglion cell layer

---

**Supplemental Figure 1.** An age-matched control retina stained with GS (red) and GFAP (green) imaged with the ELM en face demonstrated the honeycomb pattern created by the Müller cells. Very little GFAP is observed at the ELM and no processes extend beyond the ELM. This was observed consistently across the entire posterior pole. Scale bar indicates 100 µm.

**Supplemental Figure 2.** Representative control staining of Müller cell markers. (A-C) S100B (red) and GFAP (green) were both observed primarily in astrocytes in control retinas. Some isolated Müller cells also expressed these proteins. (D-F) Aquaporin 4 (red) was observed in astrocytes as well as Müller cell endfeet and perivascular processes in the inner retina. Some diffuse staining was also observed in their radial processes. No AQP4 was observed at the ELM in control retinas. GS (green) was observed throughout the entire Müller cells, including endfeet, cell bodies and radial processes. (G-I) CRALBP (red) was observed throughout the Müller cell as well as in RPE cells while GFAP (green) was confined to astrocytes. Blue label is DAPI in all images. Some background RPE labeling was observed due to insufficient quenching with Sudan black. Scale bars indicate 50 µm (same for all figures).

**Supplemental Figure 3.** Representative control staining for vimentin. (A, B) Vimentin (green) and DAPI (blue) stained control sections. Vimentin labelled the entire Müller cell body and processes. Vimentin demonstrated the linear structure of Müller cell radial processes. Scale bar indicates: 50 µm.

## References

1. Fleckenstein M, Keenan TDL, Guymer RH, et al. Age-related macular degeneration. Nat Rev Dis Primers. May 6 2021;7(1):31.

2. Edwards MM, McLeod DS, Bhutto IA, Grebe R, Duffy M, Lutty GA. Subretinal Glial Membranes in Eyes With Geographic Atrophy. Invest Ophthalmol Vis Sci. Mar 01 2017;58(3):1352–1367.

3. Edwards MM, McLeod DS, Bhutto IA, Villalonga MB, Seddon JM, Lutty GA. Idiopathic preretinal glia in aging and age-related macular degeneration. Exp Eye Res. Jul 26 2016;150:44–61.

4. Edwards MM, McLeod DS, Shen M, et al. Clinicopathologic Findings in Three Siblings With Geographic Atrophy. Invest Ophthalmol Vis Sci. Mar 1 2023;64(3):2.

5. Ramirez JM, Ramirez AI, Salazar JJ, de Hoz R, Trivino A. Changes of astrocytes in retinal ageing and age-related macular degeneration. Exp Eye Res. Nov 2001;73(5):601–615.

6. Wu KH, Madigan MC, Billson FA, Penfold PL. Differential expression of GFAP in early v late AMD: a quantitative analysis. Br J Ophthalmol. Sep 2003;87(9):1159–1166.

7. Li M, Dolz-Marco R, Huisingh C, et al. Clinicopathologic Correlation of Geographic Atrophy Secondary to Age-Related Macular Degeneration. Retina. Apr 2019;39(4):802–816.

8. Li M, Huisingh C, Messinger J, et al. HISTOLOGY OF GEOGRAPHIC ATROPHY SECONDARY TO AGE-RELATED MACULAR DEGENERATION: A Multilayer Approach. Retina. Oct 2018;38(10):1937–1953.

9. Naik P, McLeod DS, Bhutto IA, Edwards MM. Regional Alterations in Muller Cell Protein Expression in Human and a Rat Model of Geographic Atrophy. Invest Ophthalmol Vis Sci. Feb 3 2025;66(2):21.

10. Curcio CA, Messinger JD, Berlin A, et al. Fundus Autofluorescence Variation in Geographic Atrophy of Age-Related Macular Degeneration: A Clinicopathologic Correlation. Invest Ophthalmol Vis Sci. Jan 2 2025;66(1):49.

11. Dolz-Marco R, Balaratnasingam C, Messinger JD, et al. The Border of Macular Atrophy in Age-Related Macular Degeneration: A Clinicopathologic Correlation. American journal of ophthalmology. Sep 2018;193:166–177.

12. Donato R, Sorci G, Riuzzi F, et al. S100B’s double life: intracellular regulator and extracellular signal. Biochim Biophys Acta. Jun 2009;1793(6):1008–1022.

13. Donato R. Intracellular and extracellular roles of S100 proteins. Microsc Res Tech. Apr 15 2003;60(6):540–551.

14. Donato R. S100: a multigenic family of calcium-modulated proteins of the EF-hand type with intracellular and extracellular functional roles. The international journal of biochemistry & cell biology. Jul 2001;33(7):637–668.

15. Van Eldik LJ, Griffin WS. S100 beta expression in Alzheimer’s disease: relation to neuropathology in brain regions. Biochim Biophys Acta. Sep 29 1994;1223(3):398–403.

16. Sheng JG, Mrak RE, Rovnaghi CR, Kozlowska E, Van Eldik LJ, Griffin WS. Human brain S100 beta and S100 beta mRNA expression increases with age: pathogenic implications for Alzheimer’s disease. Neurobiol Aging. May–Jun 1996;17(3):359–363.

17. Sheng JG, Mrak RE, Griffin WS. S100 beta protein expression in Alzheimer disease: potential role in the pathogenesis of neuritic plaques. J Neurosci Res. Nov 1 1994;39(4):398–404.

18. Sheng JG, Mrak RE, Bales KR, et al. Overexpression of the neuritotrophic cytokine S100beta precedes the appearance of neuritic beta-amyloid plaques in APPV717F mice. J Neurochem. Jan 2000;74(1):295–301.

19. Messinger JD, Brinkmann M, Kimble JA, et al. Ex Vivo OCT-Based Multimodal Imaging of Human Donor Eyes for Research into Age-Related Macular Degeneration. J Vis Exp. May 26 2023;(195)

20. McLeod DBI, Messinger JD, Berlin A, Bijon J, Freund KB, Curcio CA, Edwards MM. Choroidal vascular changes in a case of multifocal Geographic atrophy, a clinicopathologic correlation Submitted;

21. Mj K. A formaldehyde-glutaraldehyde fixative of high osmolarity for use in electron microscopy. 1965;27:137A.

22. Tan ACS, Pilgrim MG, Fearn S, et al. Calcified nodules in retinal drusen are associated with disease progression in age-related macular degeneration. Science translational medicine. Nov 7 2018;10(466)

23. Rudolf M, Vogt SD, Curcio CA, et al. Histologic basis of variations in retinal pigment epithelium autofluorescence in eyes with geographic atrophy. Ophthalmology. Apr 2013;120(4):821–828.

24. Vogt SD, Curcio CA, Wang L, et al. Retinal pigment epithelial expression of complement regulator CD46 is altered early in the course of geographic atrophy. Exp Eye Res. Oct 2011;93(4):413–423.

25. Chen L, Li M, Messinger JD, Ferrara D, Curcio CA, Freund KB. Recognizing Atrophy and Mixed-Type Neovascularization in Age-Related Macular Degeneration Via Clinicopathologic Correlation. Transl Vis Sci Technol. Jul 2020;9(8):8.

26. Sullivan R, Penfold P, Pow DV. Neuronal migration and glial remodeling in degenerating retinas of aged rats and in nonneovascular AMD. Invest Ophthalmol Vis Sci. Feb 2003;44(2):856–865.

27. Jones BW, Pfeiffer RL, Ferrell WD, Watt CB, Tucker J, Marc RE. Retinal Remodeling and Metabolic Alterations in Human AMD. Frontiers in cellular neuroscience. 2016;10:103.

28. Jones BW, Pfeiffer RL, Ferrell WD, Watt CB, Marmor M, Marc RE. Retinal remodeling in human retinitis pigmentosa. Exp Eye Res. Mar 26 2016;

29. Jones BW, Kondo M, Terasaki H, Lin Y, McCall M, Marc RE. Retinal remodeling. Jpn J Ophthalmol. Jul 2012;56(4):289–306.

30. Edwards MM, Bonilha VL, Bhutto IA, et al. Retinal Glial and Choroidal Vascular Pathology in Donors Clinically Diagnosed With Stargardt Disease. Invest Ophthalmol Vis Sci. Jul 1 2020;61(8):27.

31. Zanzottera EC, Ach T, Huisingh C, Messinger JD, Freund KB, Curcio CA. Visualizing Retinal Pigment Epithelium Phenotypes in the Transition to Atrophy in Neovascular Age-Related Macular Degeneration. Retina. Dec 2016;36 Suppl 1:S26–S39.

32. Mrak RE, Sheng JG, Griffin WS. Correlation of astrocytic S100 beta expression with dystrophic neurites in amyloid plaques of Alzheimer’s disease. J Neuropathol Exp Neurol. Mar 1996;55(3):273–279.

33. Michetti F, Clementi ME, Di Liddo R, et al. The S100B Protein: A Multifaceted Pathogenic Factor More Than a Biomarker. Int J Mol Sci. May 31 2023;24(11)

34. Chen L, Messinger JD, Ferrara D, Freund KB, Curcio CA. Stages of Drusen-Associated Atrophy in Age-Related Macular Degeneration Visible via Histologically Validated Fundus Autofluorescence. Ophthalmology retina. Aug 2021;5(8):730–742.

35. Berlin A, Cabral D, Chen L, et al. Histology of Type 3 Macular Neovascularization and Microvascular Anomalies in Treated Age-Related Macular Degeneration: A Case Study. Ophthalmol Sci. Sep 2023;3(3):100280.

36. Cao D, Leong B, Messinger JD, et al. Hyperreflective Foci, Optical Coherence Tomography Progression Indicators in Age-Related Macular Degeneration, Include Transdifferentiated Retinal Pigment Epithelium. Invest Ophthalmol Vis Sci. Aug 2 2021;62(10):34.

37. Li M, Dolz-Marco R, Messinger JD, et al. Clinicopathologic Correlation of Anti-Vascular Endothelial Growth Factor-Treated Type 3 Neovascularization in Age-Related Macular Degeneration. Ophthalmology. Feb 2018;125(2):276–287.

38. Shu DY, Butcher E, Saint-Geniez M. EMT and EndMT: Emerging Roles in Age-Related Macular Degeneration. Int J Mol Sci. Jun 16 2020;21(12)

39. Guidry C, Medeiros NE, Curcio CA. Phenotypic variation of retinal pigment epithelium in age-related macular degeneration. Invest Ophthalmol Vis Sci. Jan 2002;43(1):267–273.

