## Supplementary figures and images for "Müller cell changes and subretinal membrane formation in an eye with multifocal geographic atrophy"

### Supplemental Fig 2

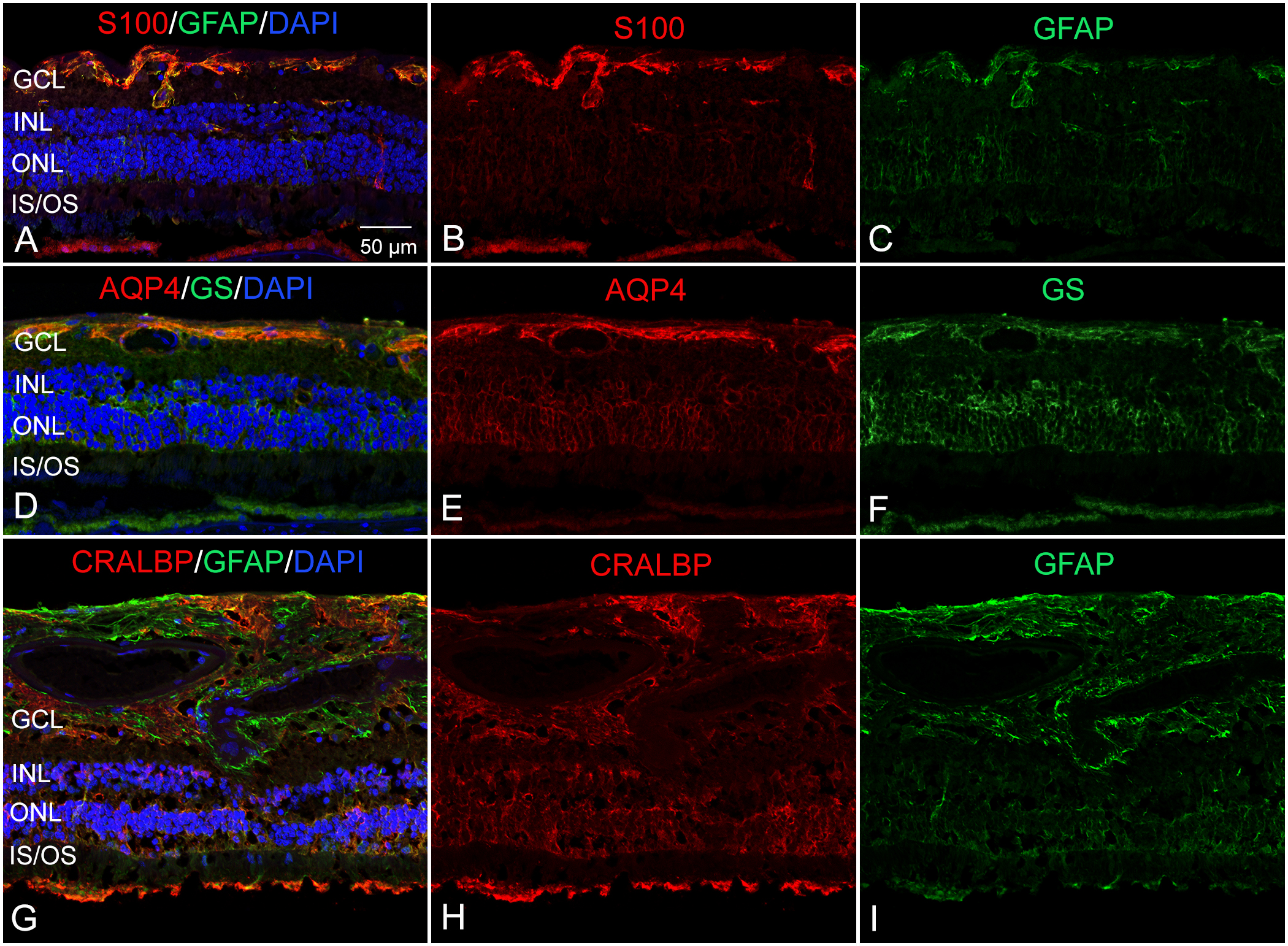

### Supplemental Fig. 1

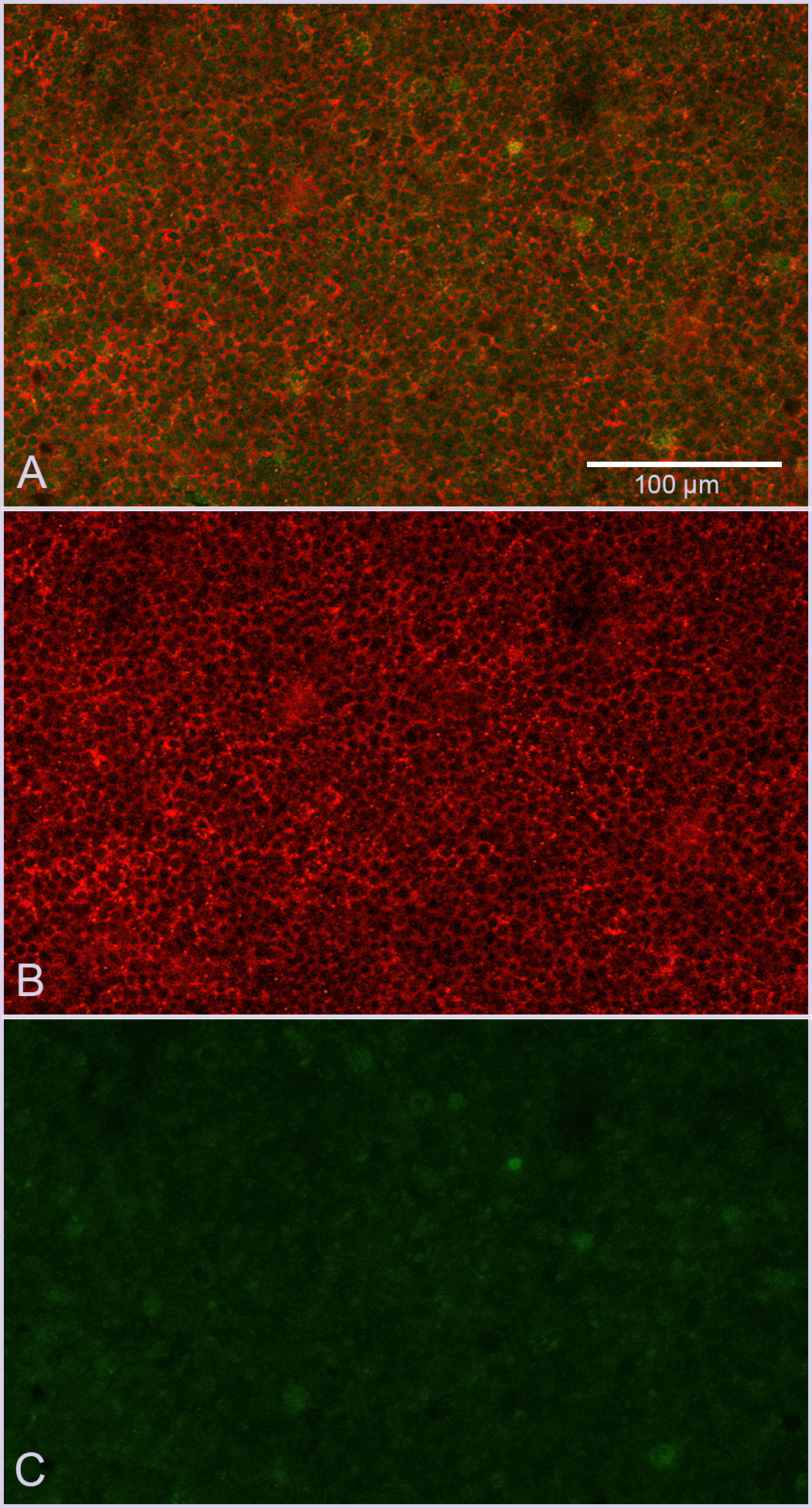

### Supplemental Fig. 3

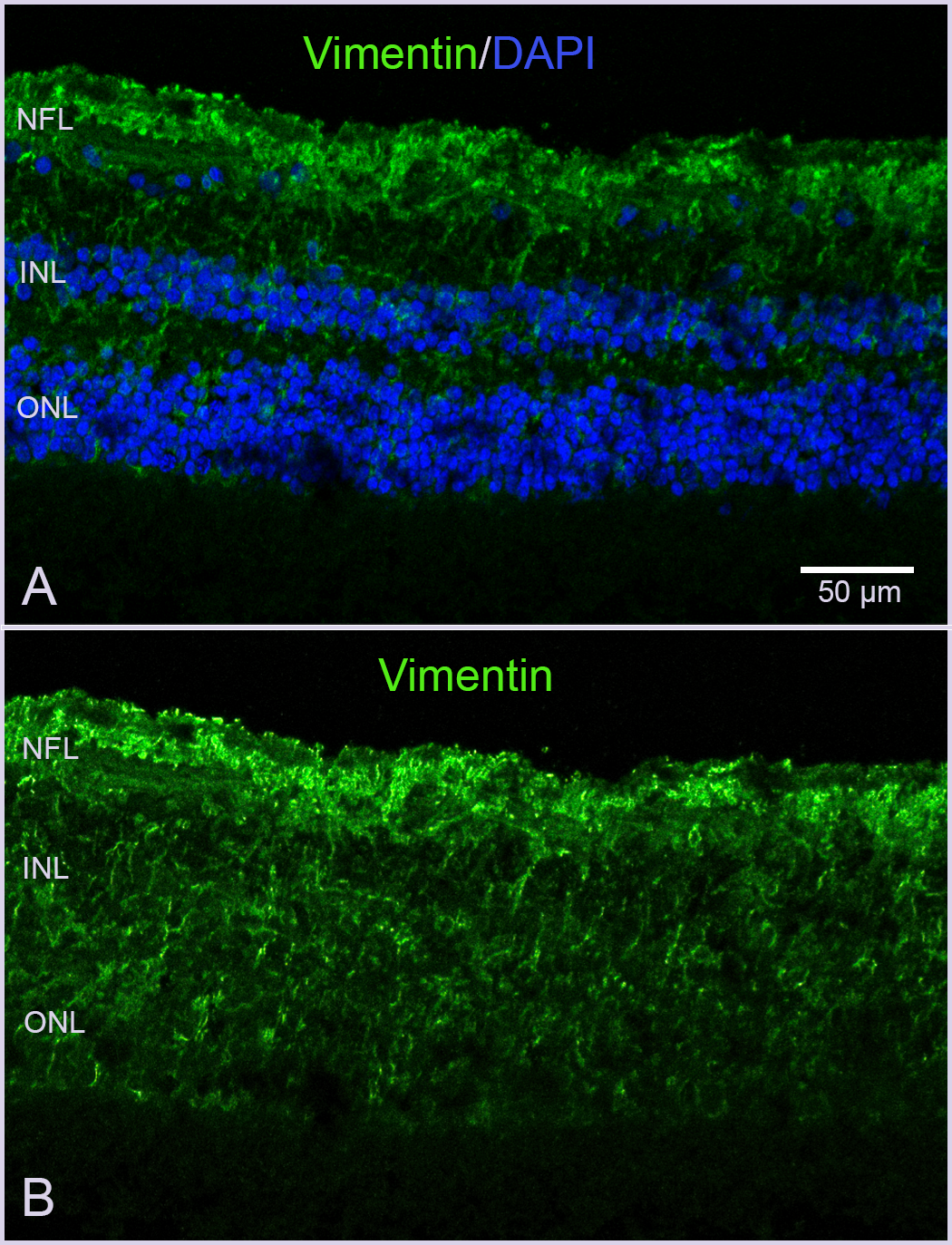
